# Genome-wide association studies of Down syndrome associated congenital heart defects

**DOI:** 10.1101/2024.09.06.24313183

**Authors:** Elizabeth R. Feldman, Yunqi Li, David J. Cutler, Tracie C. Rosser, Stephanie B. Wechsler, Lauren Sanclemente, Angela L. Rachubinski, Natalina Elliott, Paresh Vyas, Irene Roberts, Karen R. Rabin, Michael Wagner, Bruce D. Gelb, Joaquin M. Espinosa, Philip J. Lupo, Adam J. de Smith, Stephanie L. Sherman, Elizabeth J. Leslie

## Abstract

Congenital heart defects (CHDs) are the most common structural birth defect and are present in 40-50% of children born with Down syndrome (DS). To characterize the genetic architecture of DS-associated CHD, we sequenced genomes of a multiethnic group of children with DS and a CHD (n=886: atrioventricular septal defects (AVSD), n=438; atrial septal defects (ASD), n=122; ventricular septal defects (VSD), n=170; other types of CHD, n=156) and DS with a structurally normal heart (DS+NH, n=572). We performed four GWAS for common variants (MAF>0.05) comparing DS with CHD, stratified by CHD-subtype, to DS+NH controls. Although no SNP achieved genome-wide significance, multiple loci in each analysis achieved suggestive significance (p<2×10^−6^). Of these, the 1p35.1 locus (near *RBBP4*) was specifically associated with ASD risk and the 5q35.2 locus (near *MSX2*) was associated with any type of CHD. Each of the suggestive loci contained one or more plausible candidate genes expressed in the developing heart. While no SNP replicated (p<2×10^−6^) in an independent cohort of DS+CHD (DS+CHD: n=229; DS+NH: n=197), most SNPs that were suggestive in our GWASs remained suggestive when meta-analyzed with the GWASs from the replication cohort. These results build on previous work to identify genetic modifiers of DS-associated CHD.

## Introduction

Congenital heart defects (CHDs) are a heterogenous group of structural malformations that arise during embryonic heart development. They affect 8-10 of every 1000 live births and account for 25% of infant mortality (Boneva et al., 2001; Hoffman, 2013; Shuler et al., 2013), making them the most common structural birth defect in humans. CHDs have a heterogeneous etiology that includes genetic and environmental factors. One of the largest established contributors to CHD pathogenesis are chromosomal abnormalities (Hartman, Rasmussen, et al., 2011; Zaidi & Brueckner, 2017). It is estimated that 9-18% of CHD cases occur in infants with an aneuploidy, most often trisomy 21 (Abdulla, 1998; Bosi et al., 2003; Dadvand et al., 2009; Hartman, Rasmussen, et al., 2011; Schellberg et al., 2004; Zaidi & Brueckner, 2017). Trisomy 21, also known as Down syndrome (DS), is most often the consequence of nondisjunction during meiosis, resulting in an extra copy of chromosome 21. About 40-50% of infants with DS have a CHD (Freeman et al., 2008). The most common type of CHDs in this population are septal defects, the most prevalent being the atrioventricular septal defect (AVSD), followed by the ventricular septal defect (VSD) and the atrial septal defect (ASD) (Freeman et al., 2008).

AVSD is a severe form of CHD. It arises when the endocardial cushions fail to fuse during fetal valvulogenesis. The resulting defect is due to the combined effects of an ASD, VSD, and abnormalities of the mitral or tricuspid valves. In the euploid population, AVSD affects only 1 in 10,000 infants (Hartman, Riehle-Colarusso, et al., 2011), making it challenging to accumulate large cohorts needed for genetic studies. As such, little is known about the genetic architecture and etiology of AVSD. However, AVSD affects 1 in 5 infants with Down syndrome (DS) (Hartman, Riehle-Colarusso, et al., 2011), thus allowing more feasible assembly of cohorts for gene discovery. The 2000-fold increase in DS-associated AVSD compared to the euploid population suggests that aneuploidy of chromosome 21 is a principal risk factor for AVSD. However, since nearly half of infants with DS have a structurally normal heart, the extra copy of chromosome 21 alone is insufficient to cause AVSD or even abnormal cardiac development in general, suggesting that there are other factors that could modify this risk.

Many studies have previously sought to identify common variants associated with CHD and the various subtypes in euploid (Agopian et al., 2014; Cordell, Bentham, et al., 2013; Cordell, Topf, et al., 2013; Hu et al., 2013; Lahm et al., 2021) and DS populations (Ramachandran, Mulle, et al., 2015; Ramachandran, Zeng, et al., 2015; Sailani et al., 2013) with mixed results. A recent GWAS that studied over 4,000 cases with CHD and over 8,000 controls identified 20 genome-wide significant loci (Lahm et al.). As observed in prior studies, few loci could be identified when different CHD subtypes were combined. However, stratification into subtypes resulted in multiple genome-wide significant loci per group.

In DS, there has not been strong evidence that common variants contribute to CHD risk, although these studies were only powered for variants of relatively large effect. In 187 cases with DS and AVSD, ASD, or VSD and 151 controls with DS and structurally normal hearts, Sailani et al. identified no genome-wide significant loci when all CHD types were combined into a single analysis (Sailani et al.). Ramachandran et al. performed a genome-wide association study (GWAS) in 210 DS+AVSD cases and 242 DS+NH controls (Ramachandran, Zeng, et al., 2015) and also found no variants that exceeded genome wide significance, though they identified four non-chromosome 21 loci with nominal evidence of association (Ramachandran, Zeng, et al., 2015). Additionally, both Sailani et al. and Ramachandran (Ramachandran, Mulle, et al., 2015) found evidence that copy-number variants can contribute to AVSD risk in DS. In a comprehensive study of CNVs on chromosome 21, no evidence for their role in DS+AVSD was found (Rambo-Martin et al., 2018).

These studies show that phenotypic heterogeneity in CHD could negatively influence gene discovery efforts and that there may be etiologic heterogeneity and subtype-specific associations that can only be found in stratified analyses. Consistent with this, Ripoll et al., were able to distinguish the transcriptomes of lymphoblastoid cell lines established from individuals with DS with an AVSD and those with DS and an ASD or VSD, suggesting that there may be different molecular causes for different CHD subtypes (Ripoll et al., 2012). In this study, we focus on common variants (MAF > 5%) as modifiers of CHD risk in DS. Here we report the results of the largest GWAS of CHD in DS to date comparing individuals with DS and CHD (discovery: n=886; replication: n=229) to individuals with DS and structurally normal hearts (discovery: n=572; replication: n=197).

## Methods

### Study Subjects and Phenotyping

#### Discovery Cohort

Samples for this cohort were collected from four existing cohorts: the DS360 project at Emory University; the Human Trisome Project (HTP) at the Linda Crnic Institute for Down Syndrome at the University of Colorado; the Children’s Oncology Group (COG); and the Pediatric Cardiac Genomic Consortium (PCGC). Samples from individuals with Down syndrome and their unaffected parents were collected in accordance with ethical standards of human subjects’ research and with informed consent or appropriate assent, following approval by the Institutional Review Boards of the participating institutions. About 78% of samples are of predominantly European ancestry, 8% black or African American, 1% Asian ancestry and 0.5% American Indian. We collected data on the CHD phenotype from clinician and/or parent reports or medical records. Detailed CHD descriptions were collapsed into five broad categories: atrioventricular septal defect (AVSD; n=438) that included complete and incomplete AVSD, atrial septal defect (ASD; n=122), ventricular septal defect (VSD; n=170), other types of CHD (n=156), and structurally normal heart (NH; n=672). Individuals with only a patent ductus arteriosus (PDA) or patent foramen ovale (PFO) were grouped with normal heart for the purposes of this analysis.

#### Replication Cohort

The samples for this independent cohort were from the Oxford Down Syndrome Cohort Study (ODSCS) and includes 436 children with DS who were collected prospectively from birth, enrolled across 18 hospitals in the UK between 2006 and 2012, and with serial clinical and hematological data from birth to age 4 years (Roberts I, 2017). The majority (64%) of newborns are of predominantly European ancestry, with smaller numbers with African (12%) or Asian (14%) ancestry. CHD phenotypes were coded into the same five categories: AVSD (n=77), ASD (n=72), VSD (n=37), other CHD (n=14), and NH (n=201). Parents gave written informed consent in accordance with the Declaration of Helsinki. The study was approved by the Thames Valley Research Ethics Committee (06MRE12-10; NIHR portfolio no. 6362).

### Whole Genome Sequencing (WGS) and Variant Calling

For both Discovery and Replication cohorts, paired-end whole genome sequencing (WGS) was completed by the Broad Institute as part of funded NIH INCLUDE and Gabriella Miller Kids First projects on an Illumina HiSeqX with a target read depth of ∼30x coverage. The data were aligned to hg38 and harmonized by the Kids First Data Research Center via Cavatica using a custom pipeline based on GATK best practices (Van der Auwera et al., 2013).The GATK genotyping workflow included base quality score recalibration (BQSR), simultaneous calling of SNPs and indels using single-sample variant calling (HaplotypeCaller), multiple-sample joint variant calling, and finally refinement with variant quality score recalibration (VQSR) and filtering of called variants. Kids First DRC pipelines are open source and made available to the public via GitHub (Alignment workflow: https://github.com/kids-first/kf-alignment-workflow and Joint genotyping workflow: https://github.com/kids-first/kf-jointgenotyping-workflow)

### Quality Control Steps

#### Discovery Cohort

Quality control (QC) was performed on 2,394 samples and 98,049,632 variants using VCFtools v0.1.13. Variant calls with genotype quality (GQ) < 20 or depth (DP) < 10 were set to missing. Variants were then filtered to remove flagged sites (filter flag not equal to “PASS”), multiallelic sites, monomorphic sites (minor allele count (MAC) < 1) and sites with > 3% missingness, leaving 74,841,842 variants. Samples were dropped for quality metrics outside of three standard deviations from the mean for missingness, theta, transition/transversion (Ts/Tv) ratio, silent/replacement ratio, and heterozygous/homozygous ratio. Sex and family relationships were confirmed by X chromosome heterozygosity and identity-by-descent analyses in PLINK v1.9. The final analysis dataset consisted of 2,344 samples. Variants were then excluded from analysis if they had MAF < 5%, deviated from Hardy-Weinberg (p-value < 1×10^−7^), or had site missingness > 5%.

#### Replication Cohort

QC was performed on 436 samples and 51,213,158 variants using VCFtools v0.1.14 and PLINK v1.9 & v2.0. Variants were filtered to exclude non-autosomal and chromosome 21 variants, multiallelic variants, those with missingness >5%, significant differential missingness between cases and controls, variants deviating from Hardy-Weinberg, and those with MAF less than 1%. Five samples were removed from the analyses following sex checks and relationship checks (only unrelated samples were retained), and low call rates (>10% missing data). Related samples were identified if the kinship coefficient was greater than 0.177 (monozygotic twins, duplicate, first-degree relatives). Sex was inferred using X chromosome inbreeding coefficient F, where smaller than 0.25 indicated female and greater than 0.8 indicated male. An additional five samples were missing information on CHD and were not included in the GWAS. The top 20 principal components were calculated using pruned SNPs. After filtering, replication datasets included 12,601,590, 12,291,231, 12,726,561, and 12,214339 variants respectively for further AVSD, ASD, VSD, and any CHD vs. NH case-control analyses.

### Statistical Analyses

#### Discovery GWAS

We performed four within-DS case-control association tests using a logistic regression with an additive model, including sex and the first 7 principal components of ancestry as covariates in Plink v 2.00a2.3LM. The top 30 principal components were calculated in Plink v 2.00a2.3LM using a list of pruned SNPs. Each group of “cases” (DS+AVSD, n=438; DS+ASD, n=122; DS+VSD, n=170; and any DS+CHD, n=886) were contrasted with DS+NH controls (n=572). All discovery GWASs included 6,095,966 variants. Variants on chromosome 21 were excluded because genotype calls on this chromosome require special treatment due to their triploid state.

#### Replication GWAS

GWAS were performed for AVSD (n=77), ASD (n=72), VSD (n=37), and any CHD (n=229) vs. NH (n=197) using logistic regression with an additive model, including sex and first 10 principal components of ancestry as covariates in Plink v2.00a3.7LM.

#### Meta-analysis

Summary statistics from the discovery and replication GWASs were used to perform a fixed effects meta-analysis on all variants present in both the discovery and replication cohorts (Any CHD: 5,600,511 variants; AVSD: 5,595,672 variants; ASD: 5,598,098 variants; VSD: 5,594,911 variants). Meta-analysis was implemented in PLINK v 1.90b5.3.

## Results

We sought to identify common variants associated with various types of CHD in DS. First, we performed a GWAS for any DS-associated CHD by comparing individuals with DS and any type of CHD (any DS+CHD) to “controls” with DS and a structurally normal heart (DS+NH). We also performed stratified subtype-specific GWAS for AVSD, ASD, and VSD to unmask SNPs associated with a specific type of CHD that may otherwise be undetected by combining all CHD types into a single analysis. For all four discovery analyses, no SNP achieved genome wide significance (pD < 5×10^−8^), but many reached suggestive evidence of association (p_D_ < 2×10^−6^) (Supplemental Table 1). We also performed GWASs in an independent replication cohort and combined the results for the discovery and replication studies by meta-analysis. For all replication GWASs, no SNP that had suggestive evidence of association in the discovery GWASs achieved p-values with nominal significance in replication analyses (p_R_ < 2×10^−6^), but for several the direction and magnitude of effects were consistent and the p-values in the meta-analysis (p_M_) were more significant than in the discovery GWAS alone. QQ-plots are shown in Supplemental Figure 1 and a comparison of suggestive SNPs from the discovery GWASs to the replication GWASs and meta-analyses are listed in Supplemental Table 2. In the next sections, we describe the major results for each GWAS.

**Table 1:** Comparison of the suggestive SNPs from a published GWAS of CHD in Down Syndrome (Sailani et al., 2013) to SNPs in the current study.

| Published SNPs from Sailani et al. |  |  |  | Current Study |  |  |  |  |  |
| --- | --- | --- | --- | --- | --- | --- | --- | --- | --- |
| Phenotype | ID | P-VALUE | ODDS RATIO | AVSD+DS v NH+DS |  | ASD+DS v NH+DS |  | VSD+DS v NH+DS |  |
|  |  |  |  | P-VALUE | ODDS RATIO | P-VALUE | ODDS RATIO | P-VALUE | ODDS RATIO |
| CHD+DS | rs1968470 | 3.05E-06 | 2.099 | NA | NA | NA | NA | NA | NA |
| CHD+DS | rs2852749 | 3.37E-06 | 0.3576 | 6.42E-01 | 1.06 | 4.79E-01 | 0.87 | 7.77E-01 | 1.05 |
| CHD+DS | **rs1354615** | 6.00E-06 | 2.043 | 2.37E-01 | 1.12 | 4.12E-01 | 0.88 | **6.15E-03** | 1.45 |
| CHD+DS | rs2875129 | 1.09E-05 | 1.991 | 6.08E-02 | 0.84 | 5.15E-01 | 0.91 | 1.80E-01 | 0.84 |
| CHD+DS | rs9928573 | 1.26E-05 | 2.107 | 3.08E-01 | 1.11 | 7.15E-01 | 1.06 | 5.44E-01 | 1.09 |
| AVSD+DS | rs1568214 | 1.72E-06 | 2.693 | 5.94E-01 | 0.95 | 7.79E-01 | 0.96 | 6.81E-01 | 0.95 |
| AVSD+DS;<br>CHD+DS | rs2983865 | 2.08E-06 | 0.3508 | 1.37E-01 | 0.86 | 5.13E-01 | 1.11 | 2.37E-01 | 0.85 |
| AVSD+DS | rs7778237 | 2.43E-06 | 2.653 | 4.75E-01 | 0.93 | 6.22E-01 | 0.92 | 7.75E-01 | 0.96 |
| AVSD+DS | rs3011868 | 2.80E-06 | 2.666 | 1.27E-01 | 0.87 | 2.30E-01 | 0.83 | 4.07E-01 | 0.90 |
| AVSD+DS;<br>CHD+DS | **rs2983879** | 5.06E-06 | 0.3711 | **4.23E-02** | 0.82 | 7.76E-01 | 1.05 | 1.85E-01 | 0.84 |
| AVSD+DS | rs2917461 | 5.26E-06 | 2.594 | 7.82E-02 | 0.85 | 2.21E-01 | 0.83 | 5.54E-01 | 0.93 |
| AVSD+DS | rs13084784 | 5.63E-06 | 2.623 | 5.38E-01 | 1.07 | 3.49E-01 | 0.85 | 8.03E-01 | 0.96 |
| AVSD+DS | **rs2983882** | 6.21E-06 | 2.554 | **4.04E-03** | 0.77 | 3.86E-01 | 0.88 | 7.53E-02 | 0.80 |
| AVSD+DS | rs6946669 | 1.24E-05 | 2.472 | 9.84E-01 | 1.00 | 7.60E-01 | 0.95 | 8.14E-01 | 0.97 |
| AVSD+DS | rs7799573 | 1.24E-05 | 2.472 | 9.55E-01 | 1.01 | 7.76E-01 | 0.96 | 8.90E-01 | 0.98 |
| ASD+DS | rs160890 | 7.83E-08 | 8.632 | 3.42E-01 | 1.17 | 6.09E-01 | 1.16 | 5.54E-01 | 0.86 |
| ASD+DS | rs779355 | 1.40E-06 | 3.076 | NA | NA | NA | NA | NA | NA |
| ASD+DS | rs7033354 | 5.26E-06 | 0.2865 | 9.81E-01 | 1.00 | 7.74E-01 | 0.96 | 9.68E-01 | 0.99 |
| ASD+DS | **rs11102433** | 8.34E-06 | 3.157 | **1.53E-02** | 0.74 | 4.82E-01 | 0.88 | **4.86E-02** | 0.71 |
| ASD+DS | rs930655 | 8.86E-06 | 0.3418 | 4.08E-01 | 0.92 | 5.34E-01 | 1.10 | 9.36E-02 | 1.26 |
| ASD+DS | rs1672460 | 9.89E-06 | 3.831 | 7.53E-01 | 0.95 | 6.26E-01 | 1.12 | 5.91E-02 | 1.42 |
| ASD+DS | rs7789571 | 1.28E-05 | 4.95 | 7.02E-01 | 0.93 | 3.67E-01 | 1.30 | 6.37E-02 | 1.53 |
| ASD+DS | rs1704394 | 1.40E-05 | 3.135 | 6.44E-01 | 0.94 | 5.15E-01 | 1.13 | 2.10E-01 | 1.23 |
| ASD+DS | rs7498941 | 2.23E-05 | 3.353 | 5.39E-01 | 0.91 | 8.48E-01 | 0.95 | 8.94E-01 | 0.97 |
| ASD+DS | rs9850770 | 2.24E-05 | 3.082 | NA | NA | NA | NA | NA | NA |
| VSD+DS | rs1949058 | 1.32E-07 | 1.284 | 2.29E-01 | 1.12 | 6.85E-01 | 1.07 | 1.94E-01 | 0.84 |
| VSD+DS | rs10788451 | 1.83E-07 | 0.5663 | 1.41E-01 | 1.15 | 1.41E-01 | 1.15 | 1.41E-01 | 1.15 |
| VSD+DS | rs1847461 | 4.81E-07 | 1.153 | 1.99E-01 | 1.25 | 3.65E-01 | 1.26 | 2.25E-01 | 0.73 |
| VSD+DS; CHD+DS | rs17119691 | 1.04E-06 | 0.927 | 5.72E-01 | 1.09 | 2.12E-01 | 1.37 | 4.46E-01 | 1.18 |
| VSD+DS; CHD+DS | rs11621241 | 1.04E-06 | 1.038 | NA | NA | NA | NA | NA | NA |
| VSD+DS | rs10859032 | 1.40E-06 | 1.021 | 1.76E-01 | 1.28 | 3.85E-01 | 1.25 | 2.85E-01 | 0.76 |
| VSD+DS | rs2393073 | 1.68E-06 | 1.176 | 3.67E-01 | 1.13 | 2.39E-01 | 1.27 | 3.66E-01 | 0.83 |
| VSD+DS | rs10859030 | 4.90E-06 | 0.824 | 2.10E-01 | 1.22 | 7.04E-01 | 1.10 | 7.52E-02 | 0.64 |
| VSD+DS | rs545830 | 6.15E-06 | 1.131 | 7.06E-01 | 0.96 | 7.71E-01 | 1.06 | 6.38E-02 | 0.73 |
| VSD+DS | **rs3747517** | 6.85E-06 | 1.016 | **1.98E-02** | 1.28 | 8.01E-01 | 0.96 | 8.36E-01 | 0.97 |

SNPs that achieved replication in the current study with nominal significance ( $p < 0.05$ ) are highlighted in **bold**.

### DS+CHD

In the any DS+CHD discovery GWAS, we identified two regions with suggestive evidence of association: 5q11.2 and 5q35.2 (Figure 1). At 5q11.2, the lead SNP was rs11747003 (p_D_ = 1.78×10^−6^, OR = 0.58), located within an intron of *ITGA1*. Although not significant in the replication (p_R_ = 0.153, OR = 0.73), the meta-analysis p-value improved (p_M_ = 9.74×10^−7^ OR = 0.60). The lead SNP at 5q35.2 was rs76104490 (p_D_ = 7.67×10^−8^, OR = 1.97). This locus is located near *MSX2*, a homeobox protein. This SNP did not achieve suggestive significance in the replication GWAS (p_R_ = 0.625, OR = 1.12), but remained suggestive in the meta-analysis (P_m_ = 7.46×10^−7^, OR = 1.73).

**Figure 1:**
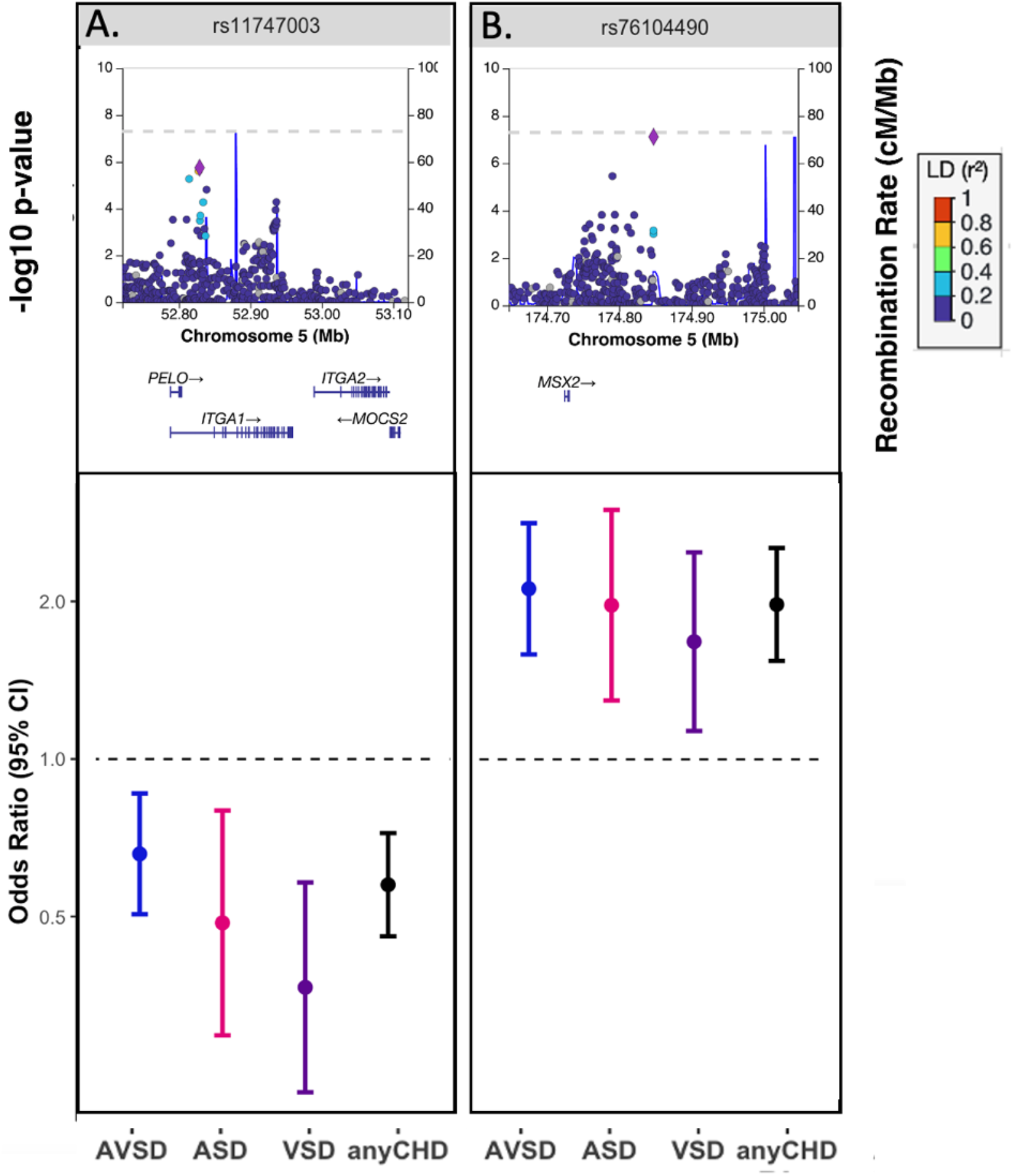
Two regions achieved suggestive significance in the any CHD+DS GWAS. The top panels show locus zoom plots for the lead SNP in each region with the left y-axis displaying the -log10(p-value) and the right y-axis displaying the recombination rate. Points are colored according to their strength of linkage disequilibrium with the lead SNP (purple diamond). The bottom panels show the odds ratio with a 95% confidence interval for the lead SNP in each region from four of the GWAS’: AVSD+DS (blue), ASD+DS (pink), VSD+DS (purple), anyCHD+DS (black).

### DS+AVSD

In addition to 5q35.2 (described above), three loci were nominated by the AVSD discovery GWAS: 12q21.2, 13q21.33, and 19q13.3 (Figure 2 A-D, top). Three SNPs were suggestive at the 12q21.2 locus (lead SNP rs7956838; p_D_ = 9.76×10^−7^, OR = 0.57), about 84 kb from *NAV3*, a neuronal regulator (Lv et al., 2022). SNPs in this region had p-values between 2.26×10^−5^ and 8.37×10^−5^ in the meta-analysis. The 13q21.33 locus (lead SNP rs9592842; p_D_ = 1.64×10^−6^, OR = 1.69) contains multiple genes predicted to be actively transcribed during heart development based on chromatin state segmentation data from human embryonic hearts (VanOudenhove et al., 2020). The lead SNP at this locus was nominally significant in the meta-analysis (p_M_ = 1.45×10^−6^, OR = 1.63). Lastly, there were nine suggestive SNPs at 19q13.3, The strongest signal was rs4807577 (p_D_ = 1.22×10^−6^, OR = 1.92), approximately 2 kb from *FSD1*, a centrosome-associated protein, and overlaps a known embryonic heart enhancer (Supplemental Figure 2) (Whyte et al., 2013).

**Figure 2:**
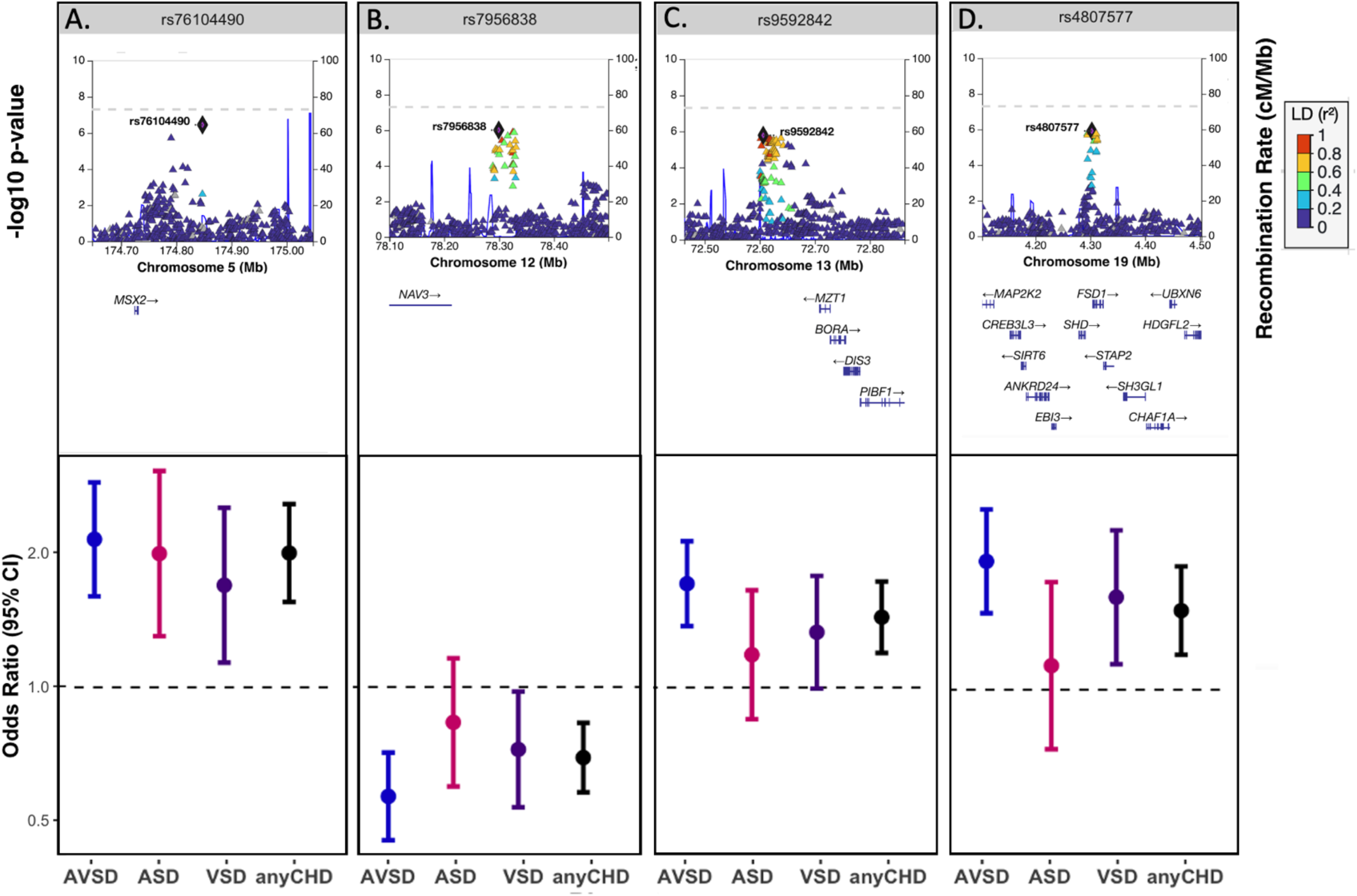
Four regions achieved suggestive significance in the AVSD+DS GWAS. The top panels show locus zoom plots for the lead SNP in each region with the left y-axis displaying the -log10(p-value) and the right y-axis displaying the recombination rate. Points are colored according to their strength of linkage disequilibrium with the lead SNP (purple diamond). The bottom panels show the odds ratio with a 95% confidence interval for the lead SNP in each region from four of the GWAS’: AVSD+DS (blue), ASD+DS (pink), VSD+DS (purple), anyCHD+DS (black).

Two additional loci (11p15.4 and 15q36.3) reached suggestive significance in the replication GWAS which did not in the discovery GWAS. The lead SNP at the 11p15.4 locus, rs2179 (p_R_ = 1.87×10^−06^, OR = 3.25), is located within an intron of *TRIM22*. The lead SNP (rs7169638) at the 15q36.3 locus approached genome-wide significance (p_R_ = 7.44×10^−8^, OR = 4.15). SNPs at this locus are in the vicinity of embryonic heart enhancers, but the nearby genes (*MEX2B, ELF1*) are not yet known to be involved in human heart conditions.

### DS+ASD

In the DS+ASD discovery, we identified six loci: 1p35.1, 1q41, 8p11.21, 10p11.21, 10q26.3, and 12q13.12 (Figure 3 A-F). Two SNPs were suggestive within 1p35.1. The lead SNP was rs75999698 (p_D_ = 3.32×10^−7^, OR = 3.15), 365 bp from *RBBP4*, a histone binding subunit. This locus was the only one to maintain suggestive significance in the meta-analysis (p_M_ = 1.87×10^−6^, OR = 2.45). The 1q41 signal was in an intergenic region between *USH2A* and *ESRRG* (lead SNP rs34506231, p_D_ = 1.98×10^−6^, OR = 0.42). At 8p11.21, there was one suggestive SNP, rs12545518 (p_D_ = 6.46×10^−7^, OR = 3.44), near *KAT6A*. At locus 10p11.21, 13 SNPs had p-value < 2×10^−6^, including lead SNP rs72794622 (p_D_ = 1.93×10^−6^, OR = 3.61). This locus is 938 kb from *FZD8*, a *Wnt* receptor expressed in the heart and associated with hypertrophic cardiomyopathy (Bergmann, 2010). Two SNPs were suggestive at the 10p26.3 locus (lead SNP rs12415529; p_D_ = 9.04×10^−7^, OR =3.09). While there are many genes at this locus, none are known to be associated with the heart in the existing literature. Finally, the 12q13.12 locus contained four suggestive SNPs (rs608233; p_D_ = 2.57×10^−7^, OR = 2.69). This locus contains many genes predicted to be actively transcribed during human heart development (VanOudenhove et al., 2020). Further, one SNP (rs632151) overlaps with a known embryonic heart enhancer (Supplemental Figure 3) (Whyte et al., 2013) and shows some evidence of being an eQTL in GTEx (Supplemental Figure 4-5) (Consortium, 2013).

**Figure 3:**
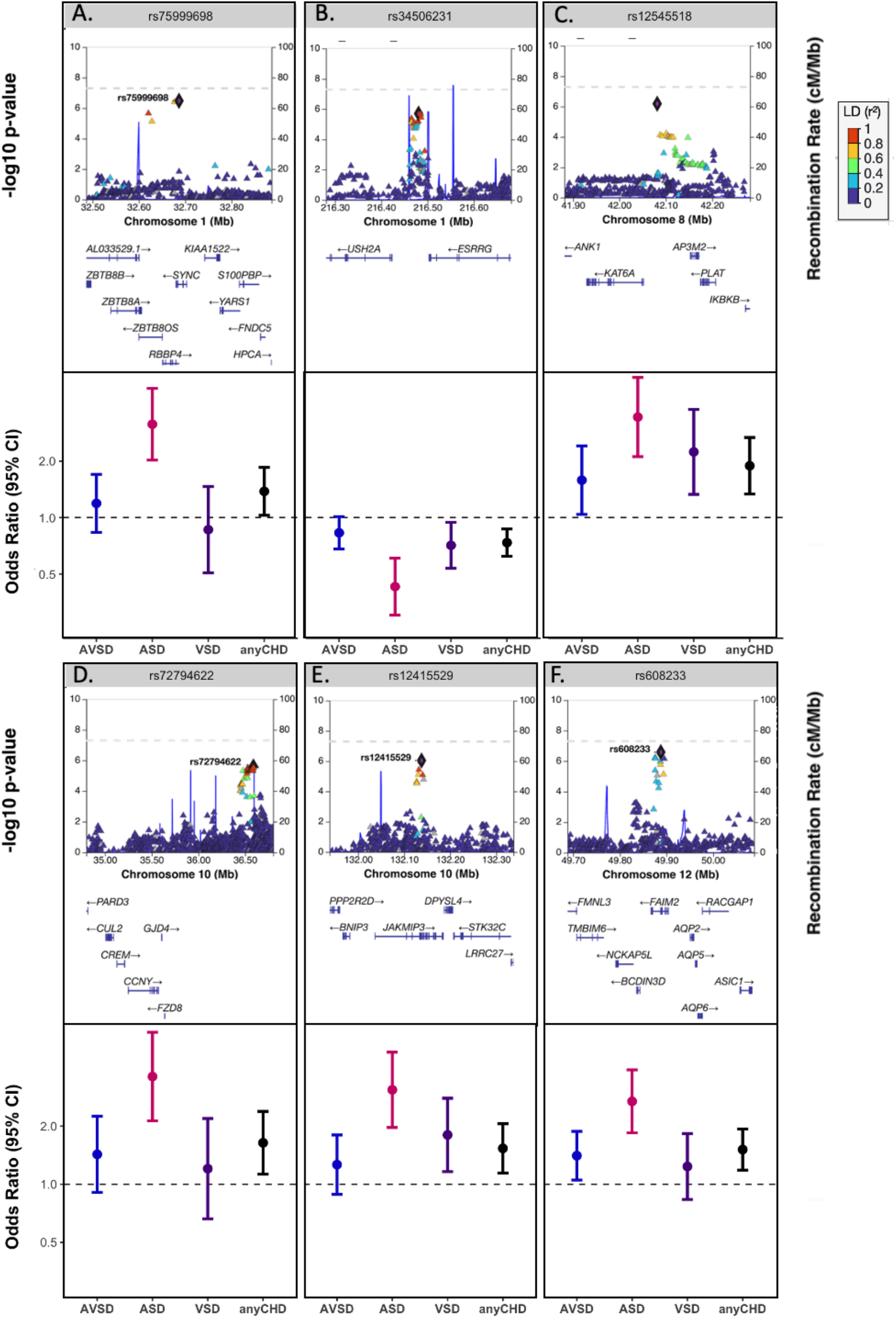
Six regions achieved suggestive significance in the ASD+DS GWAS. The top panels show locus zoom plots for the lead SNP in each region with the left y-axis displaying the -log10(p-value) and the right y-axis displaying the recombination rate. Points are colored according to their strength of linkage disequilibrium with the lead SNP (purple diamond). The bottom panels show the odds ratio with a 95% confidence interval for the lead SNP in each region from four of the GWAS’: AVSD+DS (blue), ASD+DS (pink), VSD+DS (purple), anyCHD+DS (black).

One SNP (rs1005999) at 2q12.1 in a gene desert achieved suggestive significance in the replication GWAS (p_R_ = 1.38×10^−6^, OR=3.82). SNPs at two additional loci achieved suggestive evidence of association in the meta-analysis: rs19793 at 3p26.3 (p_M_ = 1.07×10^−6^, OR = 0.47), located within *CNTN6*, and rs78087419 at 10p15.3 (p_M_ = 8.36×10^−7^, OR=3.04), located in a gene desert.

### DS+VSD

In the DS+VSD analysis, we identified four regions with suggestive evidence of association: 4q34.3, 5q33.1, 14q21.2, and 17q24.1 (Figure 4 A-D, top). At 5q33.1, two SNPs had p-values < 2×10^−6^, including the lead SNP rs9764868 (p_D_ = 8.74×10^−7^, OR = 1.98). The lead SNP is approximately 1.08 Mb from *HAND1* (Reamon-Buettner et al., 2008; Reamon-Buettner et al., 2009) and 1.05 Mb from *SAP30L (Teittinen et al*., *2012);* the former is an essential gene for cardiac morphogenesis. The lead SNPs at 4q34.3 (rs12499181; p_D_ = 1.98×10^−6^, OR = 2.07) and 14q21.2 (rs4906494; p_D_ = 1.14×0^-6^, OR = 1.95), were located within gene deserts. Only one SNP was suggestive at the 17q24.1 locus (rs72838611; p_D_ = 1.22×10^−6^, OR = 2.07), located in an intron of *PRKCA*, a regulator of cardiac contractility not known to be involved in CHD. One additional locus achieved suggestive evidence of association in the meta-analysis: 4p16.2. At this locus, there were two SNPs which had p-values < 2×10^−6^ (lead SNP rs34336375; p_M_ = 6.91×10^−7^, OR = 0.49), which is 1.32 Mb from *EVC*.

**Figure 4:**
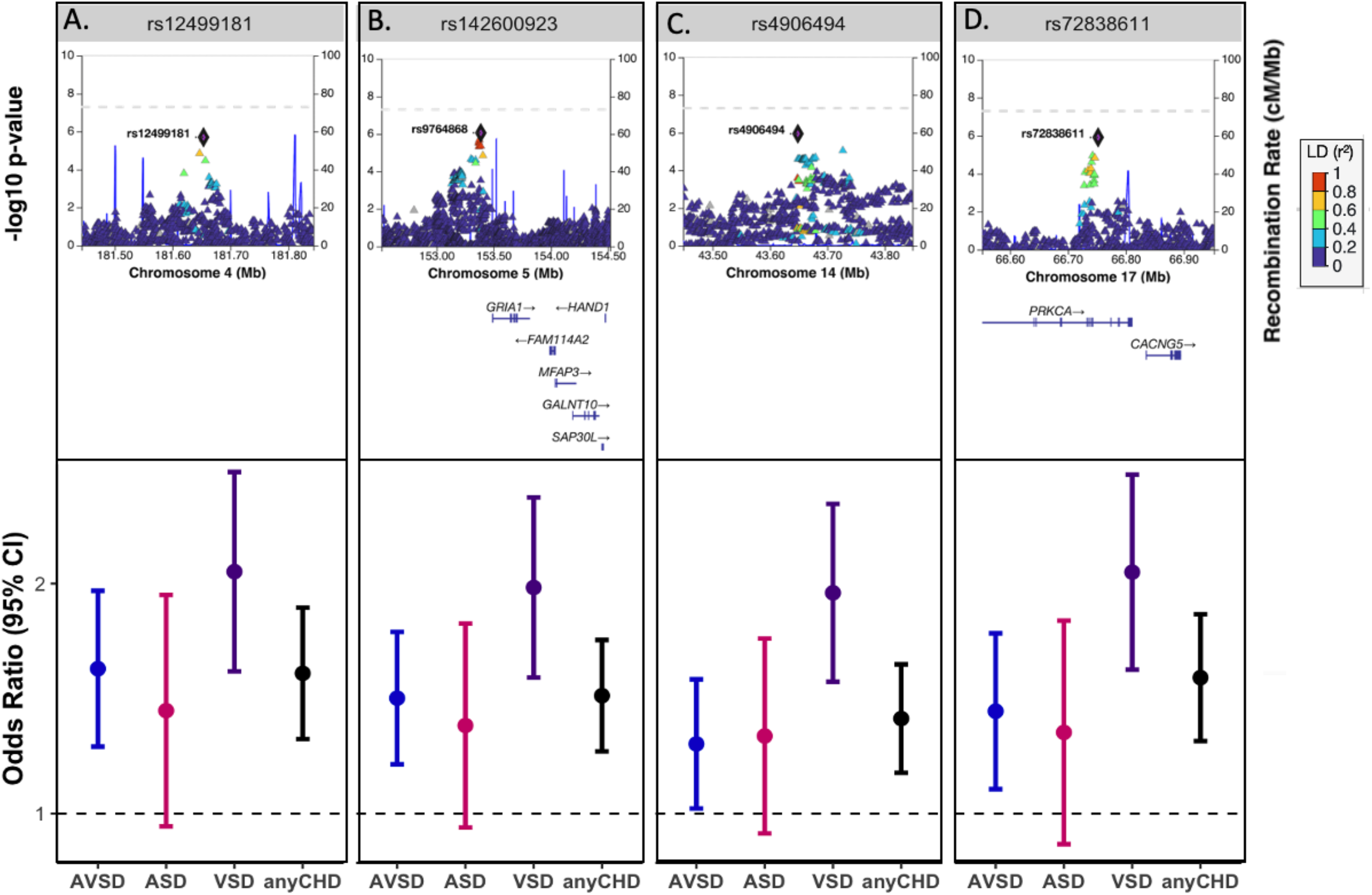
Four regions achieved suggestive significance in the VSD+DS GWAS. The top panels show locus zoom plots for the lead SNP in each region with the left y-axis displaying the -log10(p-value) and the right y-axis displaying the recombination rate. Points are colored according to their strength of linkage disequilibrium with the lead SNP (purple diamond). The bottom panels show the odds ratio with a 95% confidence interval for the lead SNP in each region from four of the GWAS’: AVSD+DS (blue), ASD+DS (pink), VSD+DS (purple), anyCHD+DS (black).

### Comparison of Effect Sizes of Discovery GWAS

To determine if any of these SNPs were specific for the CHD subtype in which they were identified, we compared the estimated odds ratio with a 95% confidence interval for the lead SNPs from any discovery subtype GWASs to the odds ratios for that SNP in each of the other three discovery GWASs. For the lead SNPs that were suggestive in the DS+AVSD discovery analysis, we found little evidence of association in the DS+ASD or DS+VSD phenotypes based on p-values. However, because the 95% CIs around the effect sizes are overlapping (Figure 2 A-D), we cannot exclude the possibility that these SNPs could also contribute a similar magnitude of risk for other types of CHD and that we have insufficient power to detect an association. We found similar results for the ASD-associated loci except for one locus. At 1p35.1, associated with DS+ASD, the confidence interval for rs75999698 in DS+ASD does not overlap that of DS+AVSD or DS+VSD (Figure 3A). The CIs for DS+AVSD and DS+VSD cross 1 (and p-values < 0.5), indicating no association. Taken together, this suggests that the 1p35.1 locus may increase risk specifically for DS+ASD. For DS+VSD discovery analysis, we observed a different pattern (Figure 4). Although all of the CIs overlapped for each subtype, the p-values were always nominally significant in AVSD (p < 0.001) but not in ASD (p < 0.01). This suggests that there may be a shared etiology between AVSD and VSD that is different from risk for ASD.

### Replication of previously published SNPs in discovery GWAS

We investigated 35 SNPs from a published GWAS of CHD and its subtypes in DS (Sailani et al., 2013) for replication in our subtype specific discovery GWASs [Table 1]. While there are other GWASs of CHD, only this study was investigated for replication since it was in an independent cohort of individuals with DS and overlapping phenotypes. Five SNPs achieved nominal significance (p < 0.05) in at least one of subtype GWASs. Two SNPs, rs2983879 (p = 5.06 × 10^−6^, OR= 0.37) and rs2983882 (p= 6.21 × 10^−6^, OR= 2.55), from the DS+AVSD GWAS in Sailani et al. (Sailani et al., 2013) replicated with nominal significance in the same phenotype in our analyses (rs2983879: p = 0.042, OR=0.82; rs2983882: p=0.004, OR=0.77). In addition, rs11102433, from the DS+ASD GWAS in Sailani et al. (Sailani et al., 2013) (p = 8.34 × 10^−6^, OR=3.15) replicated with nominal significance in DS+AVSD (p = 1.53×10^−2^, OR=0.74) and DS+VSD (p = 4.86×10^−2^, OR=0.71).

## Discussion

CHDs occur more often in children with DS than euploid children, indicating that the dosage of genes on chromosome 21 is an important risk factor for abnormal heart development. However, since not every child with DS has a CHD, we hypothesized that there are other genetic and environmental factors that modify the risk of developing a CHD in the presence of an extra copy of chromosome 21. In this study, we sought to identify common variants (MAF > 5%) outside of chromosome 21 that are modifiers of CHD risk in DS. We performed a within-DS case-control GWAS for all types of CHD. Then, we stratified the CHD cases by CHD type and performed additional GWASs of AVSD, ASD, and VSD. While no SNP achieved genome-wide significance, multiple SNPs from each discovery GWAS achieved suggestive significance. We attempted to replicate these findings in an independent cohort of DS and CHD, however most of these failed to replicate. This may be a consequence of insufficient power resulting from a small sample size, differences in race/ethnicities between the discovery and replication cohorts, heterogeneity in CHD risk, differences in CHD phenotyping, or a combination of these factors. There may be non-genetic or epigenetic factors that influence the development of CHD in DS (Mouat et al., 2023).

Although not genome-wide significant, many loci contained genes known to play key roles in heart development. These included 5q35.2, associated with all types of CHD and AVSD. The associated region contains multiple conserved and putative human cardiac enhancers approximately 115 kb downstream from *MSX2. MSX2* encodes a homeobox protein that, together with its partner MSX1, protects secondary heart field precursors against apoptosis and regulates proliferation of cardiac neural crest cells (Chen et al., 2007). In support of the critical role for MSX2 in heart development, of the 37 individuals in the DECIPHER database with a CNV in *MSX2*, ∼10% (4/37) present with an ASD, ∼10% (4/37) present with a VSD, and ∼5% (2/37) present with an abnormality of the cardiovascular system (Firth et al., 2009). Similarly, the 5q33.1 locus (associated with VSD), contained many genes in this region expressed in embryonic cardiac tissue, including *HAND1*. HAND1, with its partner HAND2, has a role in the formation of the right ventricle and aortic arches. Variants in *HAND1* have been identified in tissue samples from individuals with septal defects (AVSD, ASD, and VSD) (Reamon-Buettner et al., 2009) and hypoplastic hearts (Reamon-Buettner et al., 2008).

Multiple lines of evidence from human genetics to animal models demonstrate an important role for cilia in heart development. In an ENU screen in mice, half of the mutations identified from animals screened for congenital heart defects were in cilia related genes (Li et al., 2015). Recessive alleles in structural components of cilia and signaling machinery are associated with a variety of syndromes including CHD, laterality defects, and other cardiac abnormalities (Djenoune et al., 2022). In this study, we identified associations with several loci containing cilia genes including 13q21.33 (AVSD) and 19q13.3 (AVSD). The 13q21.33 locus contains *PIBF1*, a component of centriolar satellites (Kumar et al., 2021; Mansour et al., 2021) and the 19q13.3 locus contains *FSD1*, encoding a centrosome protein (Tu et al., 2018). Both genes result in cardiac looping defects when deleted or perturbed in animal models (Kumar et al., 2021; Liu et al., 2019).

We also identified associations near genes implicated in heart development but not yet associated with human phenotypes. An example of this is the 12q21.2 locus (AVSD), which contains *NAV3. NAV3* encodes a microtubule-binding protein that modulates neural cell migration during nervous system development (Maes et al., 2002). While *NAV3* is primarily expressed in the brain, it is also expressed in the heart, kidney, and liver (Maes et al., 2002). Although not previously associated with CHD in humans, *nav3*^-/-^ mutant zebrafish present with severe cardiac defects (Lv et al., 2022). Furthermore, expression of *nav3* in the developing zebrafish heart occurs from 24 to 48 hours post fertilization (Lv et al., 2022), when cardiac looping and the formation of the atrioventricular canals occurs (Bakkers*, 2011), suggesting a role for *nav3* in atrial-ventricle differentiation (Lv et al., 2022). Expression data based on chromHMM marks shows that *NAV3* is expressed in human cardiac tissue throughout various Carnegie stages (VanOudenhove et al., 2020).

The 1p35 locus was the only to show a clear specificity of association with one subtype. This locus was associated with ASD and contained the gene *RBBP4*, a component of the nucleosome remodeling and histone deacetylase (NuRD) complex, which interacts with known heart development gene *TBX5* (Boogerd & Evans, 2016). *RBBP4* has no reported associations with cardiac abnormalities in humans; however, it is transcribed in human embryonic cardiac tissue at all Carnegie stages (VanOudenhove et al., 2020). Notably, TBX5 is a marker of a subset of cells that ultimately becomes the atrial septum (Nadeau et al., 2010) and mutations in *TBX5* cause Holt-Oram syndrome, which frequently includes ASD or VSD among the clinical presentation (McDermott et al., 1993).

This study only focused on variants outside of chromosome 21. However, genetic variation on chromosome 21 may also contribute to CHD risk in DS. Analysis of trisomic genotypes on chromosome 21 is essential and ongoing. Similarly, this study only focused on single nucleotide polymorphisms and did not consider that CNVs may modify risk for CHD in DS.

In summary, this study identified multiple loci that are worthy of follow-up in larger cohorts of individuals with DS+CHD and also in cohorts of chromosomally typical CHD, as we have no reason yet to suspect these loci may be specific to DS. The many loci identified suggest the possibility of a significant polygenic or genetically heterogeneous risk for CHD in DS (and possibly in CHD in general). It also underscores the importance of inclusion of individuals with DS in genetic studies for CHD and the potential power of stratifying by specific type of CHD.

## Data Availability

All data analyzed in this study is available from the Kids First Data Portal and dbGaP.

## Acknowledgments

This work is a collaborative effort supported by X01-HL145686 (PJL), X01-HD117361 (JME), X01-HD110902 (JME), X01-HD107382 (JME), X01-HD107380 (AD), R01-CA249867 (KRR, PJL), R03-HL156476 (EJL, DJC), U01-HL153009 (BDG), and the Global Down Syndrome Foundation. We also thank the individuals with Down syndrome and their families for participating in this research along with the efforts of the many staff members who engaged with and collaborated with families.

## Notes

### Competing Interest Statement

The authors have declared no competing interest.

### Funding Statement

This was supported by X01-HL145686 (PJL), X01-HD117361 (JME), X01-HD110902 (JME), X01-HD107382 (JME), X01-HD107380 (AD), R01-CA249867 (KRR, PJL), R03-HL156476 (EJL, DJC), U01-HL153009 (BDG), and the Global Down Syndrome Foundation.

### Author Declarations

The IRB of Emory University, University of Colorado, Baylor College of Medicine gave ethical approval for this work. The Thames Valley Research Ethics Committee gave ethical approval for this work.

## Refrences

Abdulla, R. (1998). Perspective in pediatric cardiology. Volume 5. Genetic and environmental risk factors of major cardiovascular malformations. Pediatr Cardiol, 19(5), 435. 10.1007/s002469900346

Agopian, A. J., Mitchell, L. E., Glessner, J., Bhalla, A. D., Sewda, A., Hakonarson, H., & Goldmuntz, E. (2014). Genome-wide association study of maternal and inherited loci for conotruncal heart defects. PLoS One, 9(5), e96057. 10.1371/journal.pone.0096057

Bakkers*, J. (2011). Zebrafish as a model to study cardiac development and human cardiac disease. Cardiovasc Res.

Bergmann, M. W. (2010). WNT signaling in adult cardiac hypertrophy and remodeling: lessons learned from cardiac development. Circ Res, 107(10), 1198–1208. 10.1161/CIRCRESAHA.110.223768

Boneva, R. S., Botto, L. D., Moore, C. A., Yang, Q., Correa, A., & Erickson, J. D. (2001). Mortality associated with congenital heart defects in the United States: trends and racial disparities, 1979-1997. Circulation, 103(19), 2376–2381. 10.1161/01.cir.103.19.2376

Boogerd, C. J., & Evans, S. M. (2016). TBX5 and NuRD Divide the Heart. Dev Cell, 36(3), 242–244. 10.1016/j.devcel.2016.01.015

Bosi, G., Garani, G., Scorrano, M., Calzolari, E., & Party, I. W. (2003). Temporal variability in birth prevalence of congenital heart defects as recorded by a general birth defects registry. J Pediatr, 142(6), 690–698. 10.1067/mpd.2003.243

Chen, Y. H., Ishii, M., Sun, J., Sucov, H. M., & Maxson, R. E., Jr. (2007). Msx1 and Msx2 regulate survival of secondary heart field precursors and post-migratory proliferation of cardiac neural crest in the outflow tract. Dev Biol, 308(2), 421–437. 10.1016/j.ydbio.2007.05.037

Consortium, G. T. (2013). The Genotype-Tissue Expression (GTEx) project. Nat Genet, 45(6), 580–585. 10.1038/ng.2653

Cordell, H. J., Bentham, J., Topf, A., Zelenika, D., Heath, S., Mamasoula, C., Cosgrove, C., Blue, G., Granados-Riveron, J., Setchfield, K., Thornborough, C., Breckpot, J., Soemedi, R., Martin, R., Rahman, T. J., Hall, D., van Engelen, K., Moorman, A. F., Zwinderman, A. H., Keavney, B. D. (2013). Genome-wide association study of multiple congenital heart disease phenotypes identifies a susceptibility locus for atrial septal defect at chromosome 4p16. Nat Genet, 45(7), 822–824. 10.1038/ng.2637

Cordell, H. J., Topf, A., Mamasoula, C., Postma, A. V., Bentham, J., Zelenika, D., Heath, S., Blue, G., Cosgrove, C., Granados Riveron, J., Darlay, R., Soemedi, R., Wilson, I. J., Ayers, K. L., Rahman, T. J., Hall, D., Mulder, B. J., Zwinderman, A. H., van Engelen, K., Goodship, J. A. (2013). Genome-wide association study identifies loci on 12q24 and 13q32 associated with tetralogy of Fallot. Hum Mol Genet, 22(7), 1473–1481. 10.1093/hmg/dds552

Dadvand, P., Rankin, J., Shirley, M. D., Rushton, S., & Pless-Mulloli, T. (2009). Descriptive epidemiology of congenital heart disease in Northern England. Paediatr Perinat Epidemiol, 23(1), 58–65. 10.1111/j.1365-3016.2008.00987.x

Djenoune, L., Berg, K., Brueckner, M., & Yuan, S. (2022). A change of heart: new roles for cilia in cardiac development and disease. Nat Rev Cardiol, 19(4), 211–227. 10.1038/s41569-021-00635-z

Firth, H. V., Richards, S. M., Bevan, A. P., Clayton, S., Corpas, M., Rajan, D., Van Vooren, S., Moreau, Y., Pettett, R. M., & Carter, N. P. (2009). DECIPHER: Database of Chromosomal Imbalance and Phenotype in Humans Using Ensembl Resources. Am J Hum Genet, 84(4), 524–533. 10.1016/j.ajhg.2009.03.010

Freeman, S. B., Bean, L. H., Allen, E. G., Tinker, S. W., Locke, A. E., Druschel, C., Hobbs, C. A., Romitti, P. A., Royle, M. H., Torfs, C. P., Dooley, K. J., & Sherman, S. L. (2008). Ethnicity, sex, and the incidence of congenital heart defects: a report from the National Down Syndrome Project. Genet Med, 10(3), 173–180. 10.1097/GIM.0b013e3181634867

Hartman, R. J., Rasmussen, S. A., Botto, L. D., Riehle-Colarusso, T., Martin, C. L., Cragan, J. D., Shin, M., & Correa, A. (2011). The contribution of chromosomal abnormalities to congenital heart defects: a population-based study. Pediatr Cardiol, 32(8), 1147–1157. 10.1007/s00246-011-0034-5

Hartman, R. J., Riehle-Colarusso, T., Lin, A., Frias, J. L., Patel, S. S., Duwe, K., Correa, A., Rasmussen, S. A., & National Birth Defects Prevention, S. (2011). Descriptive study of nonsyndromic atrioventricular septal defects in the National Birth Defects Prevention Study, 1997-2005. Am J Med Genet A, 155A(3), 555–564. 10.1002/ajmg.a.33874

Hoffman, J. (2013). The global burden of congenital heart disease. Cardiovasc J Afr, 24(4), 141–145. 10.5830/CVJA-2013-028

Hu, Z., Shi, Y., Mo, X., Xu, J., Zhao, B., Lin, Y., Yang, S., Xu, Z., Dai, J., Pan, S., Da, M., Wang, X., Qian, B., Wen, Y., Wen, J., Xing, J., Guo, X., Xia, Y., Ma, H., … Shen, H. (2013). A genome-wide association study identifies two risk loci for congenital heart malformations in Han Chinese populations. Nat Genet, 45(7), 818–821. 10.1038/ng.2636

Kumar, D., Rains, A., Herranz-Perez, V., Lu, Q., Shi, X., Swaney, D. L., Stevenson, E., Krogan, N. J., Huang, B., Westlake, C., Garcia-Verdugo, J. M., Yoder, B. K., & Reiter, J. F. (2021). A ciliopathy complex builds distal appendages to initiate ciliogenesis. J Cell Biol, 220(9). 10.1083/jcb.202011133

Lahm, H., Jia, M., Dressen, M., Wirth, F., Puluca, N., Gilsbach, R., Keavney, B. D., Cleuziou, J., Beck, N., Bondareva, O., Dzilic, E., Burri, M., Konig, K. C., Ziegelmuller, J. A., Abou-Ajram, C., Neb, I., Zhang, Z., Doppler, S. A., Mastantuono, E., … Krane, M. (2021). Congenital heart disease risk loci identified by genome-wide association study in European patients. J Clin Invest, 131(2). 10.1172/JCI141837

Li, Y., Klena, N. T., Gabriel, G. C., Liu, X., Kim, A. J., Lemke, K., Chen, Y., Chatterjee, B., Devine, W., Damerla, R. R., Chang, C., Yagi, H., San Agustin, J. T., Thahir, M., Anderton, S., Lawhead, C., Vescovi, A., Pratt, H., Morgan, J., … Lo, C. W. (2015). Global genetic analysis in mice unveils central role for cilia in congenital heart disease. Nature, 521(7553), 520–524. 10.1038/nature14269

Liu, Z., Tu, H., Kang, Y., Xue, Y., Ma, D., Zhao, C., Li, H., Wang, L., & Liu, F. (2019). Primary cilia regulate hematopoietic stem and progenitor cell specification through Notch signaling in zebrafish. Nat Commun, 10(1), 1839. 10.1038/s41467-019-09403-7

Lv, F., Ge, X., Qian, P., Lu, X., Liu, D., & Chen, C. (2022). Neuron navigator 3 (NAV3) is required for heart development in zebrafish. Fish Physiol Biochem, 48(1), 173–183. 10.1007/s10695-022-01049-5

Maes, T., Barcelo, A., & Buesa, C. (2002). Neuron navigator: a human gene family with homology to unc-53, a cell guidance gene from Caenorhabditis elegans. Genomics, 80(1), 21–30. 10.1006/geno.2002.6799

Mansour, F., Boivin, F. J., Shaheed, I. B., Schueler, M., & Schmidt-Ott, K. M. (2021). The Role of Centrosome Distal Appendage Proteins (DAPs) in Nephronophthisis and Ciliogenesis. Int J Mol Sci, 22(22). 10.3390/ijms222212253

McDermott, D. A., Fong, J. C., & Basson, C. T. (1993). Holt-Oram Syndrome. In M. P. Adam, J. Feldman, G. M. Mirzaa, R. A. Pagon, S. E. Wallace, L. J. H. Bean, K. W. Gripp, & A. Amemiya (Eds.), GeneReviews((R)). https://www.ncbi.nlm.nih.gov/pubmed/20301290

Mouat, J. S., Li, S., Myint, S. S., Laufer, B. I., Lupo, P. J., Schraw, J. M., Woodhouse, J. P., de Smith, A. J., & LaSalle, J. M. (2023). Epigenomic signature of major congenital heart defects in newborns with Down syndrome. Hum Genomics, 17(1), 92. 10.1186/s40246-023-00540-1

Nadeau, M., Georges, R. O., Laforest, B., Yamak, A., Lefebvre, C., Beauregard, J., Paradis, P., Bruneau, B. G., Andelfinger, G., & Nemer, M. (2010). An endocardial pathway involving Tbx5, Gata4, and Nos3 required for atrial septum formation. Proc Natl Acad Sci U S A, 107(45), 19356–19361. 10.1073/pnas.0914888107

Ramachandran, D., Mulle, J. G., Locke, A. E., Bean, L. J., Rosser, T. C., Bose, P., Dooley, K. J., Cua, C. L., Capone, G. T., Reeves, R. H., Maslen, C. L., Cutler, D. J., Sherman, S. L., & Zwick, M. E. (2015). Contribution of copy-number variation to Down syndrome-associated atrioventricular septal defects. Genet Med, 17(7), 554–560. 10.1038/gim.2014.144

Ramachandran, D., Zeng, Z., Locke, A. E., Mulle, J. G., Bean, L. J., Rosser, T. C., Dooley, K. J., Cua, C. L., Capone, G. T., Reeves, R. H., Maslen, C. L., Cutler, D. J., Feingold, E., Sherman, S. L., & Zwick, M. E. (2015). Genome-Wide Association Study of Down Syndrome-Associated Atrioventricular Septal Defects. G3 (Bethesda), 5(10), 1961–1971. 10.1534/g3.115.019943

Rambo-Martin, B. L., Mulle, J. G., Cutler, D. J., Bean, L. J. H., Rosser, T. C., Dooley, K. J., Cua, C., Capone, G., Maslen, C. L., Reeves, R. H., Sherman, S. L., & Zwick, M. E. (2018). Analysis of Copy Number Variants on Chromosome 21 in Down Syndrome-Associated Congenital Heart Defects. G3 (Bethesda), 8(1), 105–111. 10.1534/g3.117.300366

Reamon-Buettner, S. M., Ciribilli, Y., Inga, A., & Borlak, J. (2008). A loss-of-function mutation in the binding domain of HAND1 predicts hypoplasia of the human hearts. Hum Mol Genet, 17(10), 1397–1405. 10.1093/hmg/ddn027

Reamon-Buettner, S. M., Ciribilli, Y., Traverso, I., Kuhls, B., Inga, A., & Borlak, J. (2009). A functional genetic study identifies HAND1 mutations in septation defects of the human heart. Hum Mol Genet, 18(19), 3567–3578. 10.1093/hmg/ddp305

Ripoll, C., Rivals, I., Ait Yahya-Graison, E., Dauphinot, L., Paly, E., Mircher, C., Ravel, A., Grattau, Y., Blehaut, H., Megarbane, A., Dembour, G., de Freminville, B., Touraine, R., Creau, N., Potier, M. C., & Delabar, J. M. (2012). Molecular signatures of cardiac defects in Down syndrome lymphoblastoid cell lines suggest altered ciliome and Hedgehog pathways. PLoS One, 7(8), e41616. 10.1371/journal.pone.0041616

Roberts I B.N., Chaussade A, et al. (2017). Mature Analysis of the Prospective Oxford Down Syndrome (DS) Cohort Study: Timing and Clinical Impact of Preleukemic GATA1 Mutations and Lessons for Management of Newborns with DS. blood, 130.

Sailani, M. R., Makrythanasis, P., Valsesia, A., Santoni, F. A., Deutsch, S., Popadin, K., Borel, C., Migliavacca, E., Sharp, A. J., Duriaux Sail, G., Falconnet, E., Rabionet, K., Serra-Juhe, C., Vicari, S., Laux, D., Grattau, Y., Dembour, G., Megarbane, A., Touraine, R., … Antonarakis, S. E. (2013). The complex SNP and CNV genetic architecture of the increased risk of congenital heart defects in Down syndrome. Genome Res, 23(9), 1410–1421. 10.1101/gr.147991.112

Schellberg, R., Schwanitz, G., Gravinghoff, L., Kallenberg, R., Trost, D., Raff, R., & Wiebe, W. (2004). New trends in chromosomal investigation in children with cardiovascular malformations. Cardiol Young, 14(6), 622–629. 10.1017/S1047951104006079

Shuler, C. O., Black, G. B., & Jerrell, J. M. (2013). Population-based treated prevalence of congenital heart disease in a pediatric cohort. Pediatr Cardiol, 34(3), 606–611. 10.1007/s00246-012-0505-3

Teittinen, K. J., Gronroos, T., Parikka, M., Junttila, S., Uusimaki, A., Laiho, A., Korkeamaki, H., Kurppa, K., Turpeinen, H., Pesu, M., Gyenesei, A., Ramet, M., & Lohi, O. (2012). SAP30L (Sin3A-associated protein 30-like) is involved in regulation of cardiac development and hematopoiesis in zebrafish embryos. J Cell Biochem, 113(12), 3843–3852. 10.1002/jcb.24298

Tu, H. Q., Qin, X. H., Liu, Z. B., Song, Z. Q., Hu, H. B., Zhang, Y. C., Chang, Y., Wu, M., Huang, Y., Bai, Y. F., Wang, G., Han, Q. Y., Li, A. L., Zhou, T., Liu, F., Zhang, X. M., & Li, H. Y. (2018). Microtubule asters anchored by FSD1 control axoneme assembly and ciliogenesis. Nat Commun, 9(1), 5277. 10.1038/s41467-018-07664-2

Van der Auwera, G. A., Carneiro, M. O., Hartl, C., Poplin, R., Del Angel, G., Levy-Moonshine, A., Jordan, T., Shakir, K., Roazen, D., Thibault, J., Banks, E., Garimella, K. V., Altshuler, D., Gabriel, S., & DePristo, M. A. (2013). From FastQ data to high confidence variant calls: the Genome Analysis Toolkit best practices pipeline. Curr Protoc Bioinformatics, 43(1110), 11 10 11–11 10 33. 10.1002/0471250953.bi1110s43

VanOudenhove, J., Yankee, T. N., Wilderman, A., & Cotney, J. (2020). Epigenomic and Transcriptomic Dynamics During Human Heart Organogenesis. Circ Res, 127(9), e184–e209. 10.1161/CIRCRESAHA.120.316704

Whyte, W. A., Orlando, D. A., Hnisz, D., Abraham, B. J., Lin, C. Y., Kagey, M. H., Rahl, P. B., Lee, T. I., & Young, R. A. (2013). Master transcription factors and mediator establish super-enhancers at key cell identity genes. Cell, 153(2), 307–319. 10.1016/j.cell.2013.03.035

Zaidi, S., & Brueckner, M. (2017). Genetics and Genomics of Congenital Heart Disease. Circ Res, 120(6), 923–940. 10.1161/CIRCRESAHA.116.309140

