## Supplemental for "Genome-wide association studies of Down syndrome associated congenital heart defects"

**Supplemental Table 1: All suggestive SNPs from Discovery GWASs**

| | | | | | any CHD+DS | | AVSD+DS | | ASD+DS | | VSD+DS | |
|---|---|---|---|---|---|---|---|---|---|---|---|---|
| **LOCUS** | **CHR** | **POS** | **ID** | **REF/ALT*** | **P** | **OR** | **P** | **OR** | **P** | **OR** | **P** | **OR** |
| 1p35.1 | 1 | 32676829 | rs34621149 | G/T | 3.11E-02 | 1.36 | 3.13E-01 | 1.19 | **3.92E-07** | **3.08** | 6.47E-01 | 0.89 |
| | | 32686571 | rs75999698 | G/A | 3.19E-02 | 1.38 | 3.38E-01 | 1.19 | **3.32E-07** | **3.15** | 5.82E-01 | 0.86 |
| 1q41 | 1 | 216480519 | rs34506231 | AT/A | 3.15E-04 | 0.74 | 6.59E-02 | 0.83 | **1.98E-06** | **0.43** | 1.78E-02 | 0.71 |
| 4q34,3 | 4 | 181653471 | rs12499181 | G/T | 2.84E-05 | 1.53 | 2.70E-04 | 1.55 | 8.13E-02 | 1.36 | **1.98E-06** | **2.07** |
| 5q11.2 | 5 | 52829053 | rs11747003 | A/C | **1.78E-06** | **0.57** | 2.10E-03 | 0.66 | 4.26E-03 | 0.49 | 1.99E-05 | 0.37 |
| 5q33.1 | 5 | 153375118 | rs11957080 | G/A | 5.72E-05 | 1.41 | 8.44E-04 | 1.40 | 1.22E-01 | 1.28 | **1.99E-06** | **1.93** |
| | | 153386992 | rs142600923 | A/G | 3.35E-05 | 1.43 | 6.32E-04 | 1.42 | 9.07E-02 | 1.30 | **8.74E-07** | **1.98** |
| 5q35.2 | 5 | 174790580 | rs10050415 | C/T | 3.47E-06 | 1.87 | **1.92E-06** | **2.10** | 5.08E-01 | 1.18 | 2.77E-04 | 2.09 |
| | | 174847461 | rs76104490 | C/T | **7.67E-08** | **1.97** | **3.55E-07** | **2.12** | 1.52E-03 | 1.97 | 9.77E-03 | 1.68 |
| 8p11.21 | 8 | 42081349 | rs12545518 | C/T | 3.32E-04 | 1.89 | 3.23E-02 | 1.58 | **6.46E-07** | **3.44** | 2.43E-03 | 2.24 |
| 10p11.21 | 10 | 36582876 | rs72794616 | T/C | 9.80E-03 | 1.64 | 1.25E-01 | 1.43 | **1.93E-06** | **3.61** | 5.43E-01 | 1.20 |
| | | 36584210 | rs78718821 | A/G | 1.00E-02 | 1.64 | 1.27E-01 | 1.43 | **1.95E-06** | **3.61** | 5.46E-01 | 1.20 |
| | | 36585058 | rs12570806 | C/T | 9.80E-03 | 1.64 | 1.25E-01 | 1.43 | **1.93E-06** | **3.61** | 5.43E-01 | 1.20 |
| | | 36585614 | rs12573127 | A/G | 9.80E-03 | 1.64 | 1.25E-01 | 1.43 | **1.93E-06** | **3.61** | 5.43E-01 | 1.20 |
| | | 36585981 | rs12569478 | G/A | 9.80E-03 | 1.64 | 1.25E-01 | 1.43 | **1.93E-06** | **3.61** | 5.43E-01 | 1.20 |
| | | 36586234 | rs58403209 | T/TG | 9.55E-03 | 1.64 | 1.21E-01 | 1.43 | **1.93E-06** | **3.61** | 5.43E-01 | 1.20 |
| | | 36586431 | rs61214442 | G/T | 9.80E-03 | 1.64 | 1.25E-01 | 1.43 | **1.93E-06** | **3.61** | 5.43E-01 | 1.20 |
| | | 36587018 | rs201450144 | G/GAAAGA | 1.00E-02 | 1.64 | 1.26E-01 | 1.43 | **1.98E-06** | **3.61** | 5.46E-01 | 1.20 |

| | | | | | | | | | | | | |
|---|---|---|---|---|---|---|---|---|---|---|---|---|
| | | 36588162 | rs56333929 | A/G | 9.80E-03 | 1.64 | 1.25E-01 | 1.43 | **1.93E-06** | **3.61** | 5.43E-01 | 1.20 |
| | | 36588273 | rs56387875 | G/A | 9.80E-03 | 1.64 | 1.25E-01 | 1.43 | **1.93E-06** | **3.61** | 5.43E-01 | 1.20 |
| | | 36589141 | rs57900399 | C/T | 1.01E-02 | 1.64 | 1.26E-01 | 1.43 | **1.98E-06** | **3.61** | 5.48E-01 | 1.20 |
| | | 36589142 | rs59992697 | A/G | 9.80E-03 | 1.64 | 1.25E-01 | 1.43 | **1.93E-06** | **3.61** | 5.43E-01 | 1.20 |
| | | 36589514 | rs72794622 | T/A | 9.80E-03 | 1.64 | 1.25E-01 | 1.43 | **1.93E-06** | **3.61** | 5.43E-01 | 1.20 |
| 10q26.3 | 10 | 132136232 | rs112106400 | C/A | 1.25E-02 | 1.44 | 3.36E-01 | 1.18 | **1.07E-06** | **3.03** | 1.71E-02 | 1.69 |
| | | 132137596 | rs12415529 | C/T | 4.44E-03 | 1.53 | 1.97E-01 | 1.26 | **9.04E-07** | **3.09** | 8.54E-03 | 1.80 |

| | | | | | | | | | | | | |
|---|---|---|---|---|---|---|---|---|---|---|---|---|
| 12q13.12 | 12 | 49875962 | rs836966 | T/C | 3.07E-03 | 1.45 | 1.38E-01 | 1.25 | **5.99E-07** | **2.81** | 2.54E-01 | 1.27 |
| | | 49875979 | rs836965 | A/G | 2.90E-03 | 1.46 | 1.35E-01 | 1.25 | **5.64E-07** | **2.82** | 2.48E-01 | 1.27 |
| | | 49876112 | rs836964 | A/G | 3.54E-03 | 1.44 | 1.48E-01 | 1.24 | **6.83E-07** | **2.79** | 2.62E-01 | 1.26 |
| | | 49886933 | rs611282 | G/A | 9.56E-03 | 1.45 | 1.15E-01 | 1.30 | **1.05E-06** | **2.85** | 3.59E-01 | 1.23 |
| | | 49887266 | rs612674 | C/A | 7.07E-03 | 1.43 | 9.29E-02 | 1.30 | **1.70E-06** | **2.64** | 6.20E-01 | 1.11 |
| | | 49887864 | rs608233 | G/A | 9.64E-04 | 1.51 | 2.18E-02 | 1.41 | **2.57E-07** | **2.69** | 2.92E-01 | 1.24 |
| | | 49895440 | rs632151 | G/A | 1.06E-02 | 1.42 | 2.96E-01 | 1.19 | **7.03E-07** | **2.86** | 3.19E-01 | 1.25 |
| 12q21.2 | 12 | 78299308 | rs7956838 | A/T | 6.65E-05 | 0.70 | **9.76E-07** | **0.57** | 2.80E-01 | 0.84 | 3.43E-02 | 0.73 |
| | | 78325122 | rs7961876 | G/A | 2.03E-05 | 0.68 | **1.27E-06** | **0.57** | 3.42E-01 | 0.85 | 4.21E-02 | 0.73 |
| | | 78327445 | rs10777907 | C/T | 3.74E-05 | 0.64 | **1.34E-06** | **0.51** | 2.80E-01 | 0.81 | 9.13E-02 | 0.74 |
| 13q21.33 | 13 | 72606225 | rs9592842 | G/A | 1.14E-04 | 1.43 | **1.64E-06** | **1.69** | 3.18E-01 | 1.18 | 5.48E-02 | 1.32 |

| | | | | | | | | | | | | |
|---|---|---|---|---|---|---|---|---|---|---|---|---|
| 14q21.2 | 14 | 43648793 | rs4906494 | C/T | 5.99E-04 | 1.33 | 3.47E-02 | 1.23 | 1.19E-01 | 1.26 | **1.14E-06** | **1.95** |
| 17q24.1 | 17 | 66752111 | rs72838611 | G/T | 2.69E-05 | 1.51 | 1.01E-02 | 1.36 | 1.55E-01 | 1.28 | **1.22E-06** | **2.07** |
| 19q13.3 | 19 | 4294626 | rs58237134 | C/A | 2.86E-03 | 1.35 | **1.72E-06** | **1.76** | 9.20E-01 | 0.98 | 3.98E-02 | 1.39 |
| | | 4300409 | rs7408794 | T/G | 7.48E-04 | 1.46 | **1.40E-06** | **1.88** | 7.95E-01 | 1.06 | 1.92E-02 | 1.49 |
| | | 4302796 | rs4807577 | G/A | 4.49E-04 | 1.49 | **1.22E-06** | **1.92** | 5.72E-01 | 1.13 | 6.75E-03 | 1.60 |
| | | 4310725 | rs11883395 | T/G | 1.01E-03 | 1.46 | **1.75E-06** | **1.90** | 8.64E-01 | 1.04 | 1.35E-02 | 1.54 |
| | | 4310799 | rs36112398 | C/A | 9.91E-04 | 1.46 | **1.75E-06** | **1.90** | 8.64E-01 | 1.04 | 1.19E-02 | 1.55 |

*Odds ratio is reported for the underlined (effect) allele.
Suggestive SNPs ($p<2x10^{6}$) are **bolded** and lead SNPs are highlighted in gray. For all SNPs, the P-value and OR for that SNP in all analyses is shown for all GWASs

**Supplemental Table 2: Comparison of suggestive SNPS in discovery, replication, and meta GWASs**

| | | | | | Discovery | | Replication | | Meta | |
|---|---|---|---|---|---|---|---|---|---|---|
| Phenotype | CHR | POS | ID | REF/ALT | P | OR | P | OR | P | OR |
| DS+CHD | 5 | 52814387 | rs1391985 | C/T | 5.37E-06 | 0.563429 | 0.042168 | 0.591516 | 6.36E-07 | 0.5687 |
| | 5 | 52827565 | rs66778208 | C/T | 2.49E-06 | 0.555733 | 0.0327564 | 0.569843 | 2.35E-07 | 0.5583 |
| | 5 | 52829053 | rs11747003 | A/C | 1.78E-06 | 0.575282 | 0.153768 | 0.727304 | 9.74E-07 | 0.6046 |
| | 5 | 174847461 | rs76104490 | C/T | 7.67E-08 | 1.97466 | 0.625287 | 1.11955 | 7.46E-07 | 1.7325 |
| | 14 | 104668054 | rs12432277 | G/C | 0.0181793 | 0.820707 | 1.99E-06 | 0.481066 | 1.39E-05 | 0.7266 |
| DS+AVSD | 5 | 174790580 | rs10050415 | C/T | 1.92E-06 | 2.1048 | 0.102373 | 1.61813 | 6.64E-07 | 1.9867 |
| | 5 | 174847461 | rs76104490 | C/T | 3.55E-07 | 2.1164 | 0.441968 | 1.28015 | 7.47E-07 | 1.9395 |
| | 11 | 5701877 | rs2179 | G/C | 0.635449 | 1.05306 | 1.87E-06 | 3.25429 | 0.01848 | 1.2651 |
| | 12 | 78299308 | rs7956838 | A/T | 9.76E-07 | 0.574281 | 0.23813 | 1.33005 | 8.37E-05 | 0.668 |
| | 12 | 78325122 | rs7961876 | G/A | 1.27E-06 | 0.568021 | 0.728454 | 1.09022 | 2.26E-05 | 0.639 |
| | 12 | 78327445 | rs10777907 | C/T | 1.34E-06 | 0.508194 | 0.862663 | 1.04893 | 2.31E-05 | 0.5894 |
| | 13 | 72606225 | rs9592842 | G/A | 1.64E-06 | 1.69267 | 0.291563 | 1.30693 | 1.45E-06 | 1.625 |
| | 13 | 72638425 | rs972434 | A/T | 2.41E-06 | 1.72261 | 0.157383 | 1.47029 | 9.89E-07 | 1.6817 |
| | 15 | 81923413 | rs35138517 | A/C | 0.822277 | 0.972463 | 6.74E-07 | 4.16624 | 0.07711 | 1.2234 |
| | 15 | 81930489 | rs7177009 | A/G | 0.743127 | 1.04082 | 1.46E-06 | 3.90845 | 0.02722 | 1.2809 |
| | 15 | 81933243 | rs7180317 | G/T | 0.980959 | 0.997163 | 1.08E-06 | 3.84234 | 0.05619 | 1.2321 |
| | 15 | 81941749 | rs2047086 | C/T | 0.982849 | 1.00259 | 1.93E-06 | 3.79163 | 0.05752 | 1.2336 |
| | 15 | 81943348 | rs7169638 | C/T | 0.42852 | 1.09046 | 7.44E-08 | 4.3675 | 0.006332 | 1.3196 |
| | 15 | 81948719 | rs2867580 | A/G | 0.828104 | 1.0251 | 3.44E-07 | 4.14807 | 0.03305 | 1.2526 |
| | 19 | 4294626 | rs58237134 | C/A | 1.72E-06 | 1.7647 | 0.955052 | 0.986718 | 2.11E-05 | 1.5707 |
| | 19 | 4300409 | rs7408794 | T/G | 1.40E-06 | 1.88307 | 0.24105 | 0.709083 | 8.63E-05 | 1.6001 |
| | 19 | 4302796 | rs4807577 | G/A | 1.22E-06 | 1.91841 | 0.259889 | 0.718794 | 8.10E-05 | 1.6179 |
| | 19 | 4310725 | rs11883395 | T/G | 1.75E-06 | 1.89691 | 0.940839 | 1.02128 | 1.33E-05 | 1.6946 |
| | 19 | 4310799 | rs36112398 | C/A | 1.75E-06 | 1.89691 | 0.940839 | 1.02128 | 1.33E-05 | 1.6946 |
| DS+ASD | 1 | 32676829 | rs34621149 | G/T | 3.92E-07 | 3.08084 | 0.382713 | 1.36428 | 1.87E-06 | 2.4529 |
| | 1 | 32686571 | rs75999698 | G/A | 3.32E-07 | 3.1457 | 0.364119 | 1.39166 | 1.43E-06 | 2.5128 |
| | 1 | 216480519 | rs34506231 | AT/A | 1.98E-06 | 0.428232 | 0.954347 | 0.986823 | 0.0001429 | 0.5842 |
| | 2 | 104907333 | rs1005999 | C/T | 0.0862581 | 1.30808 | 1.38E-06 | 3.81931 | 0.0001106 | 1.6941 |
| | 3 | 1388226 | rs149793 | C/G | 0.00039874 | 0.5154 | 0.00053394 | 0.388021 | 1.07E-06 | 0.4707 |

| | | | | | | | | | | |
|---|---|---|---|---|---|---|---|---|---|---|
| | 8 | 42081349 | rs12545518 | C/T | 6.46E-07 | 3.43594 | 0.797567 | 1.11363 | 1.01E-05 | 2.5666 |
| | 10 | 2515328 | rs78087419 | A/G | 0.00016433 | 2.76463 | 0.00118819 | 3.83332 | 8.36E-07 | 3.0471 |
| | 10 | 36582876 | rs72794616 | T/C | 1.93E-06 | 3.61471 | 0.419829 | 0.697614 | 0.0002556 | 2.3269 |
| | 10 | 36584210 | rs78718821 | A/G | 1.95E-06 | 3.6101 | 0.419829 | 0.697614 | 0.0002589 | 2.3249 |
| | 10 | 36585058 | rs12570806 | C/T | 1.93E-06 | 3.61471 | 0.419829 | 0.697614 | 0.0002556 | 2.3269 |
| | 10 | 36585614 | rs12573127 | A/G | 1.93E-06 | 3.61471 | 0.419829 | 0.697614 | 0.0002556 | 2.3269 |
| | 10 | 36585981 | rs12569478 | G/A | 1.93E-06 | 3.61471 | 0.419829 | 0.697614 | 0.0002556 | 2.3269 |
| | 10 | 36586234 | rs58403209 | T/TG | 1.93E-06 | 3.61471 | 0.419829 | 0.697614 | 0.0002556 | 2.3269 |
| | 10 | 36586431 | rs61214442 | G/T | 1.93E-06 | 3.61471 | 0.419829 | 0.697614 | 0.0002556 | 2.3269 |
| | 10 | 36587018 | rs201450144 | G/GAAAGA | 1.98E-06 | 3.60821 | 0.419829 | 0.697614 | 0.0002594 | 2.3244 |
| | 10 | 36588162 | rs56333929 | A/G | 1.93E-06 | 3.61471 | 0.419829 | 0.697614 | 0.0002556 | 2.3269 |
| | 10 | 36588273 | rs56387875 | G/A | 1.93E-06 | 3.61471 | 0.419829 | 0.697614 | 0.0002556 | 2.3269 |
| | 10 | 36589141 | rs57900399 | C/T | 1.98E-06 | 3.60745 | 0.419829 | 0.697614 | 0.0002598 | 2.3241 |
| | 10 | 36589142 | rs59992697 | A/G | 1.93E-06 | 3.61471 | 0.419829 | 0.697614 | 0.0002556 | 2.3269 |
| | 10 | 36589514 | rs72794622 | T/A | 1.93E-06 | 3.61471 | 0.419829 | 0.697614 | 0.0002556 | 2.3269 |
| | 10 | 132136232 | rs112106400 | C/A | 1.07E-06 | 3.03296 | 0.912721 | 1.0393 | 3.24E-05 | 2.2115 |
| | 10 | 132137596 | rs12415529 | C/T | 9.04E-07 | 3.08613 | 0.684801 | 1.15576 | 1.36E-05 | 2.3149 |
| | 12 | 49875962 | rs836966 | T/C | 5.99E-07 | 2.81126 | 0.661756 | 1.13852 | 1.41E-05 | 2.0904 |
| | 12 | 49875979 | rs836965 | A/G | 5.64E-07 | 2.81903 | 0.661756 | 1.13852 | 1.35E-05 | 2.094 |
| | 12 | 49876112 | rs836964 | A/G | 6.83E-07 | 2.79467 | 0.527079 | 1.20654 | 9.19E-06 | 2.1234 |
| | 12 | 49886933 | rs611282 | G/A | 1.05E-06 | 2.85433 | 0.755328 | 0.898709 | 7.17E-05 | 2.0602 |
| | 12 | 49887266 | rs612674 | C/A | 1.70E-06 | 2.64272 | 0.404824 | 0.754536 | 0.0002392 | 1.8956 |
| | 12 | 49887864 | rs608233 | G/A | 2.57E-07 | 2.68805 | 0.777594 | 0.912276 | 1.76E-05 | 2.033 |
| | 12 | 49895440 | rs632151 | G/A | 7.03E-07 | 2.86437 | 0.558428 | 1.19393 | 1.09E-05 | 2.1476 |
| DS+VSD | 4 | 5946066 | rs13121232 | A/C | 2.74E-05 | 0.520945 | 0.00431071 | 0.352835 | 6.45E-07 | 4.91E-01 |
| | 4 | 5950704 | rs34336375 | AC/A | 3.53E-05 | 0.528623 | 0.0030005 | 0.337502 | 6.91E-07 | 4.94E-01 |
| | 4 | 181653471 | rs12499181 | G/T | 1.98E-06 | 2.07322 | 0.603608 | 1.16614 | 8.16E-06 | 1.8356 |
| | 5 | 153375118 | rs11957080 | G/A | 1.99E-06 | 1.92935 | 0.0922605 | 0.595476 | 0.0002643 | 1.5841 |
| | 5 | 153386992 | rs142600923 | A/G | 8.74E-07 | 1.97649 | 0.0922605 | 0.595476 | 0.0001588 | 1.6134 |
| | 14 | 43648793 | rs4906494 | C/T | 1.14E-06 | 1.94509 | 0.536977 | 1.19032 | 3.35E-06 | 1.77E+00 |
| | 17 | 66752111 | rs72838611 | G/T | 1.22E-06 | 2.07013 | 0.338389 | 0.693808 | 3.09E-05 | 1.79E+00 |

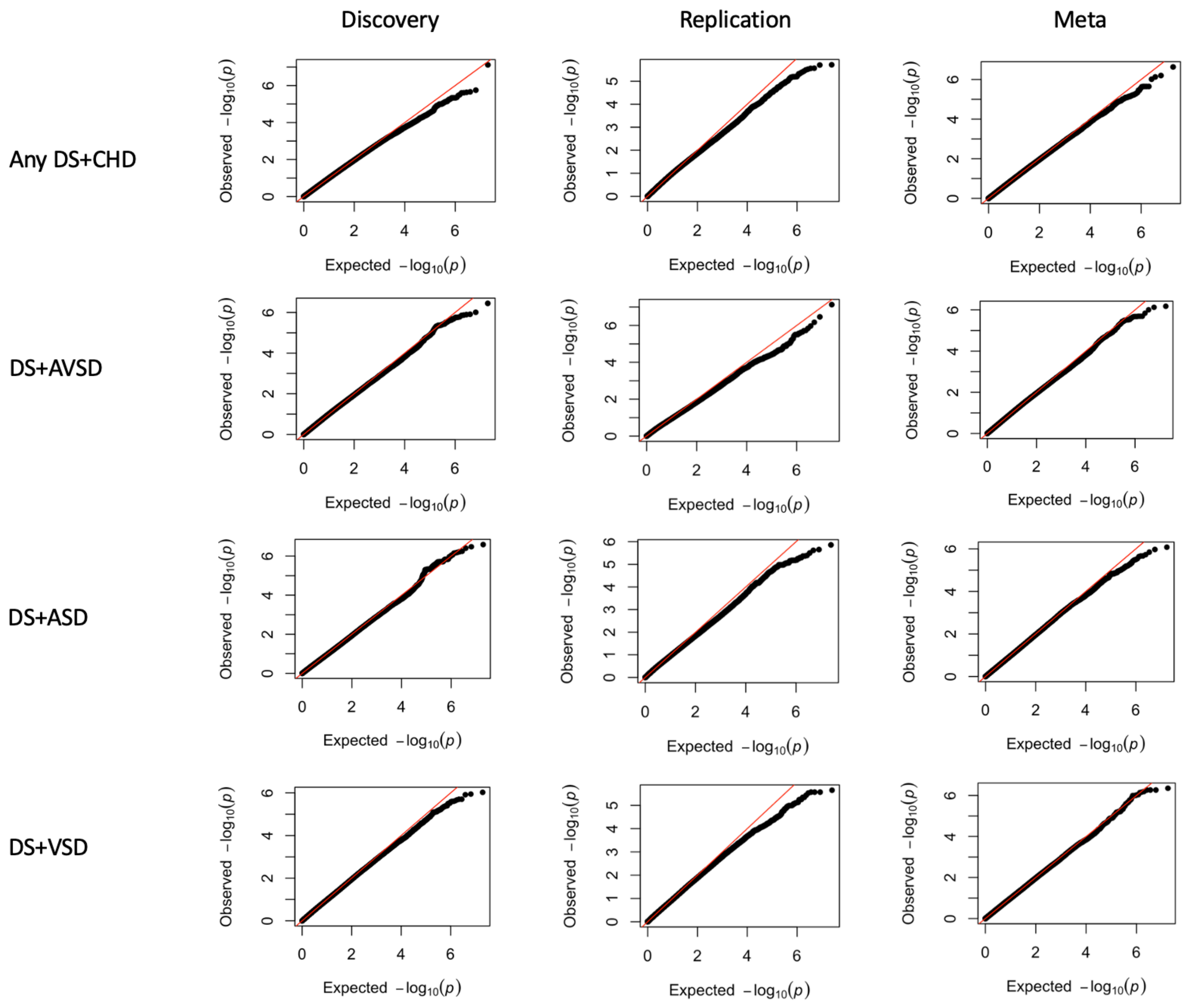


*Supplemental Figure 1:* **qq plots for each GWAS**. The columns indicate if the GWAS was the discovery, replication or meta-analysis and the rows represent the phenotype of the GWAS.

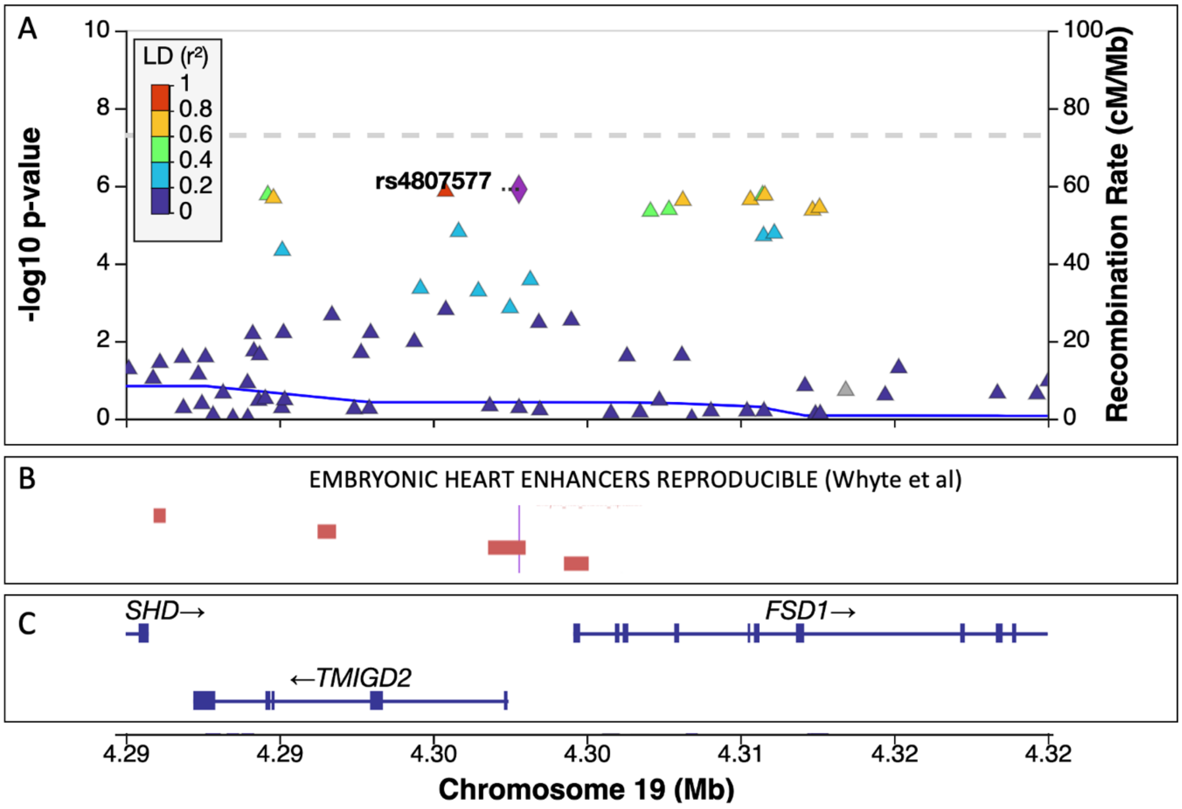


*Supplemental Figure 2:* **Lead SNP from the AVSD+DS GWAS overlaps an embryonic heart enhancer.** (A) Locus zoom plot of 19q13.3. The purple diamond indicates the SNP which overlaps the embryonic heart enhancer. Points are colored according to their strength of linkage disequilibrium with the overlapping SNP. (B) The region of known embryonic heart enhancers. The thin purple line illustrates where the overlapping SNP is located. (C) Positions of known genes in the region.

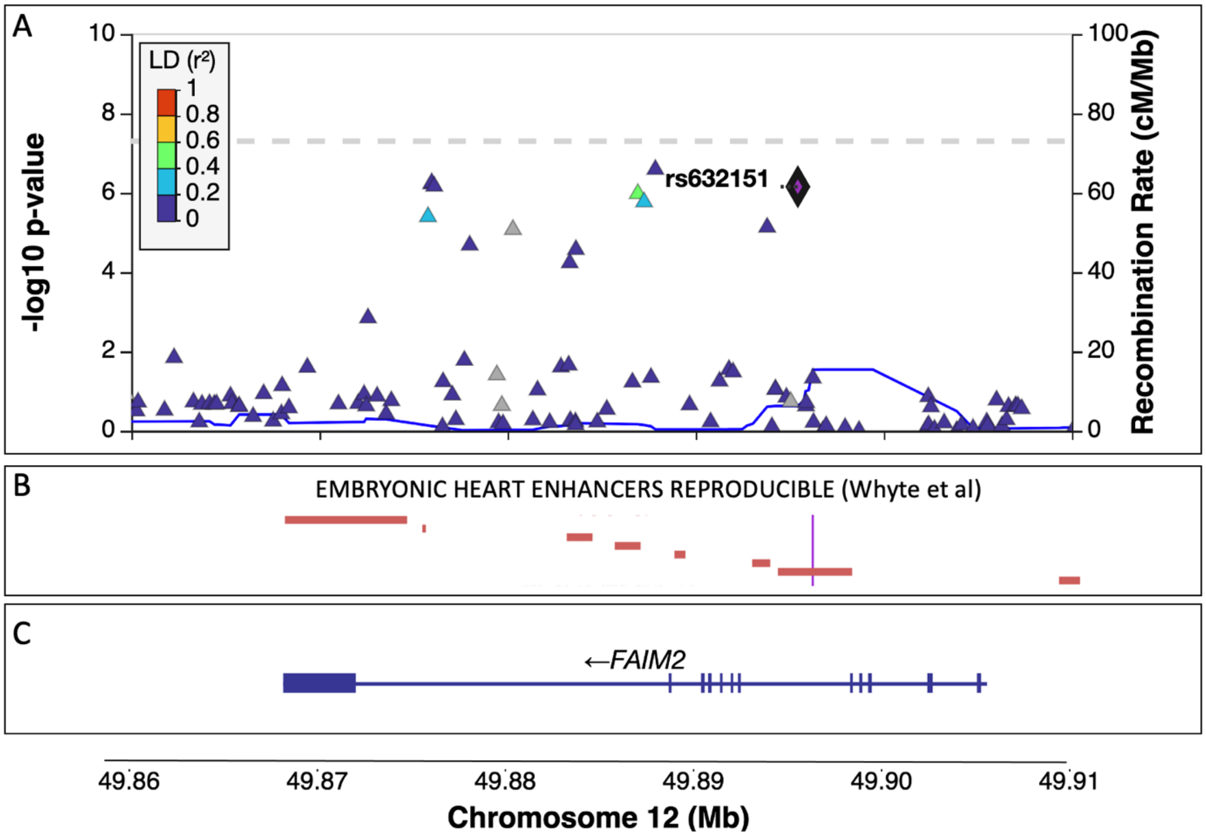


*Supplemental Figure 3:* **SNP from the ASD+DS GWAS overlaps an embryonic heart enhancer.** (A) Locus zoom plot of 12q13.12. The purple diamond indicates the SNP which overlaps the embryonic heart enhancer. Points are colored according to their strength of linkage disequilibrium with the overlapping SNP. (B) The region of known embryonic heart enhancers. The thin purple line illustrates where the overlapping SNP is located. (C) Positions of known genes in the region.

| Tissue | Samples | NES | p-value | m-value |
|---|---|---|---|---|
| Minor Salivary Gland | 144 | 0.368 | 0.02 | 0.964 |
| Cells - EBV-transformed lymphocytes | 147 | 0.323 | 1.8e-3 | 1.00 |
| Brain - Cerebellar Hemisphere | 175 | 0.317 | 0.01 | 0.981 |
| Brain - Spinal cord (cervical c-1) | 126 | 0.304 | 0.05 | 0.969 |
| Brain - Anterior cingulate cortex (BA24) | 147 | 0.273 | 0.1 | 0.926 |
| Heart - Left Ventricle | 386 | 0.252 | 0.009 | 0.995 |
| Brain - Cortex | 205 | 0.243 | 0.07 | 0.954 |
| Brain - Substantia nigra | 114 | 0.229 | 0.2 | 0.987 |
| Brain - Hypothalamus | 170 | 0.183 | 0.2 | 0.927 |
| Cells - Cultured fibroblasts | 483 | 0.176 | 1.4e-5 | 1.00 |
| Brain - Hippocampus | 165 | 0.172 | 0.2 | 0.959 |
| Stomach | 324 | 0.155 | 0.03 | 0.991 |
| Adipose - Subcutaneous | 581 | 0.147 | 0.03 | 0.993 |
| Muscle - Skeletal | 706 | 0.146 | 2.5e-3 | 0.995 |
| Whole Blood | 670 | 0.145 | 4.9e-4 | 1.00 |
| Adrenal Gland | 233 | 0.145 | 0.1 | 0.974 |
| Colon - Transverse | 368 | 0.139 | 0.07 | 0.976 |
| Esophagus - Gastroesophageal Junction | 330 | 0.136 | 0.04 | 0.994 |
| Colon - Sigmoid | 318 | 0.129 | 0.1 | 0.973 |
| Testis | 322 | 0.125 | 0.2 | 0.929 |
| Brain - Amygdala | 129 | 0.124 | 0.5 | 0.928 |
| Heart - Atrial Appendage | 372 | 0.120 | 0.04 | 0.997 |
| Breast - Mammary Tissue | 396 | 0.110 | 0.09 | 0.984 |
| Esophagus - Muscularis | 465 | 0.109 | 0.09 | 0.974 |
| Thyroid | 574 | 0.100 | 0.03 | 0.978 |
| Esophagus - Mucosa | 497 | 0.0994 | 0.08 | 0.983 |
| Uterus | 129 | 0.0958 | 0.4 | 0.917 |
| Spleen | 227 | 0.0885 | 0.4 | 0.968 |
| Pituitary | 237 | 0.0849 | 0.2 | 0.960 |
| Lung | 515 | 0.0842 | 0.08 | 0.959 |
| Skin - Sun Exposed (Lower leg) | 605 | 0.0841 | 0.08 | 0.968 |
| Brain - Nucleus accumbens (basal ganglia) | 202 | 0.0777 | 0.5 | 0.933 |
| Brain - Putamen (basal ganglia) | 170 | 0.0776 | 0.6 | 0.918 |
| Small Intestine - Terminal Ileum | 174 | 0.0774 | 0.5 | 0.946 |
| Vagina | 141 | 0.0766 | 0.5 | 0.912 |
| Pancreas | 305 | 0.0454 | 0.5 | 0.910 |
| Brain - Cerebellum | 209 | 0.0436 | 0.7 | 0.882 |
| Artery - Coronary | 213 | 0.0409 | 0.7 | 0.894 |
| Kidney - Cortex | 73 | 0.0309 | 0.9 | 0.927 |
| Brain - Frontal Cortex (BA9) | 175 | 0.0301 | 0.8 | 0.928 |
| Nerve - Tibial | 532 | 0.0284 | 0.7 | 0.779 |
| Artery - Tibial | 584 | 0.0263 | 0.7 | 0.781 |
| Ovary | 167 | 0.0261 | 0.8 | 0.843 |
| Brain - Caudate (basal ganglia) | 194 | 0.0182 | 0.9 | 0.901 |
| Skin - Not Sun Exposed (Suprapubic) | 517 | 0.0172 | 0.8 | 0.799 |
| Prostate | 221 | 0.0160 | 0.9 | 0.805 |
| Adipose - Visceral (Omentum) | 469 | 0.0141 | 0.8 | 0.543 |
| Liver | 208 | 0.00649 | 1 | 0.837 |
| Artery - Aorta | 387 | -0.00927 | 0.9 | 0.661 |

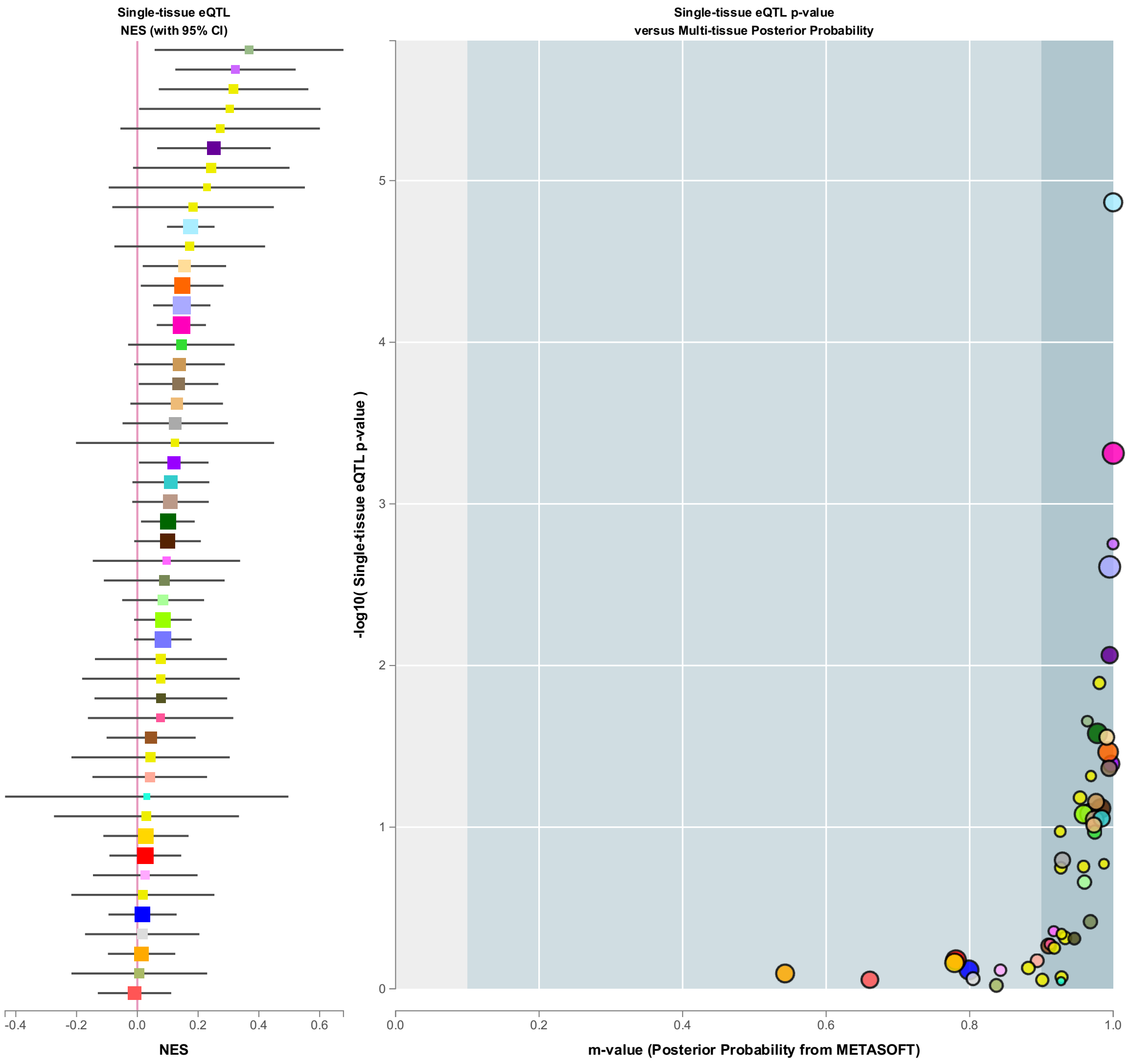


Supplemental Figure 4: **Multi-tissue eQTL Comparison. ENSG000006666.5 BCDIN3D and chr12:49895449:G:A.** Image generated in https://www.gtexportal.org/home/snp/rs632151#eqtl-block

| Tissue | Samples | NES | p-value | m-value |
|---|---|---|---|---|
| Testis | 322 | 0.152 | 4.7e-4 | 0.00 |
| Brain - Cerebellar Hemisphere | 175 | 0.135 | 0.1 | 0.0860 |
| Small Intestine - Terminal Ileum | 174 | 0.128 | 0.1 | 0.0690 |
| Minor Salivary Gland | 144 | 0.128 | 0.3 | 0.0940 |
| Heart - Atrial Appendage | 372 | 0.127 | 0.02 | 0.00 |
| Muscle - Skeletal | 706 | 0.124 | 0.03 | 0.00800 |
| Prostate | 221 | 0.114 | 0.2 | 0.0770 |
| Breast - Mammary Tissue | 396 | 0.104 | 0.2 | 0.0300 |
| Heart - Left Ventricle | 386 | 0.0839 | 0.2 | 0.0200 |
| Brain - Putamen (basal ganglia) | 170 | 0.0797 | 0.1 | 0.0130 |
| Uterus | 129 | 0.0721 | 0.7 | 0.128 |
| Kidney - Cortex | 73 | 0.0697 | 0.8 | 0.199 |
| Thyroid | 574 | 0.0663 | 0.3 | 0.0280 |
| Brain - Cerebellum | 209 | 0.0561 | 0.5 | 0.101 |
| Vagina | 141 | 0.0549 | 0.6 | 0.123 |
| Adipose - Subcutaneous | 581 | 0.0456 | 0.5 | 0.0510 |
| Brain - Caudate (basal ganglia) | 194 | 0.0376 | 0.5 | 0.0300 |
| Brain - Hippocampus | 165 | 0.0372 | 0.3 | 0.00800 |
| Pancreas | 305 | 0.0305 | 0.7 | 0.169 |
| Brain - Anterior cingulate cortex (BA24) | 147 | 0.0198 | 0.7 | 0.0470 |
| Esophagus - Mucosa | 497 | 0.0177 | 0.8 | 0.0740 |
| Colon - Transverse | 368 | 0.0173 | 0.6 | 0.0190 |
| Stomach | 324 | 0.00576 | 0.9 | 0.0860 |
| Adipose - Visceral (Omentum) | 469 | 0.00478 | 0.9 | 0.0820 |
| Artery - Tibial | 584 | 0.000593 | 1 | 0.136 |
| Pituitary | 237 | 0.000350 | 1 | 0.0970 |
| Brain - Hypothalamus | 170 | -0.00560 | 0.9 | 0.119 |
| Skin - Not Sun Exposed (Suprapubic) | 517 | -0.0157 | 0.8 | 0.133 |
| Brain - Frontal Cortex (BA9) | 175 | -0.0180 | 0.7 | 0.152 |
| Brain - Amygdala | 129 | -0.0324 | 0.6 | 0.165 |
| Esophagus - Gastroesophageal Junction | 330 | -0.0394 | 0.6 | 0.245 |
| Colon - Sigmoid | 318 | -0.0516 | 0.3 | 0.224 |
| Esophagus - Muscularis | 465 | -0.0544 | 0.3 | 0.252 |
| Whole Blood | 670 | -0.0557 | 0.4 | 0.280 |
| Liver | 208 | -0.0569 | 0.7 | 0.280 |
| Brain - Substantia nigra | 114 | -0.0591 | 0.4 | 0.267 |
| Brain - Nucleus accumbens (basal ganglia) | 202 | -0.0650 | 0.1 | 0.400 |
| Ovary | 167 | -0.0715 | 0.4 | 0.300 |
| Brain - Cortex | 205 | -0.0750 | 0.09 | 0.472 |
| Artery - Coronary | 213 | -0.0767 | 0.4 | 0.242 |
| Brain - Spinal cord (cervical c-1) | 126 | -0.0862 | 0.3 | 0.366 |
| Nerve - Tibial | 532 | -0.101 | 5.3e-5 | 0.989 |
| Adrenal Gland | 233 | -0.107 | 0.2 | 0.372 |
| Artery - Aorta | 387 | -0.118 | 0.1 | 0.499 |
| Lung | 515 | -0.118 | 0.05 | 0.551 |
| Spleen | 227 | -0.137 | 0.2 | 0.324 |
| Cells - Cultured fibroblasts | 483 | -0.164 | 0.01 | 0.719 |
| Skin - Sun Exposed (Lower leg) | 605 | -0.184 | 3.3e-3 | 0.851 |
| Cells - EBV-transformed lymphocytes | 147 | -0.470 | 0.02 | 0.462 |

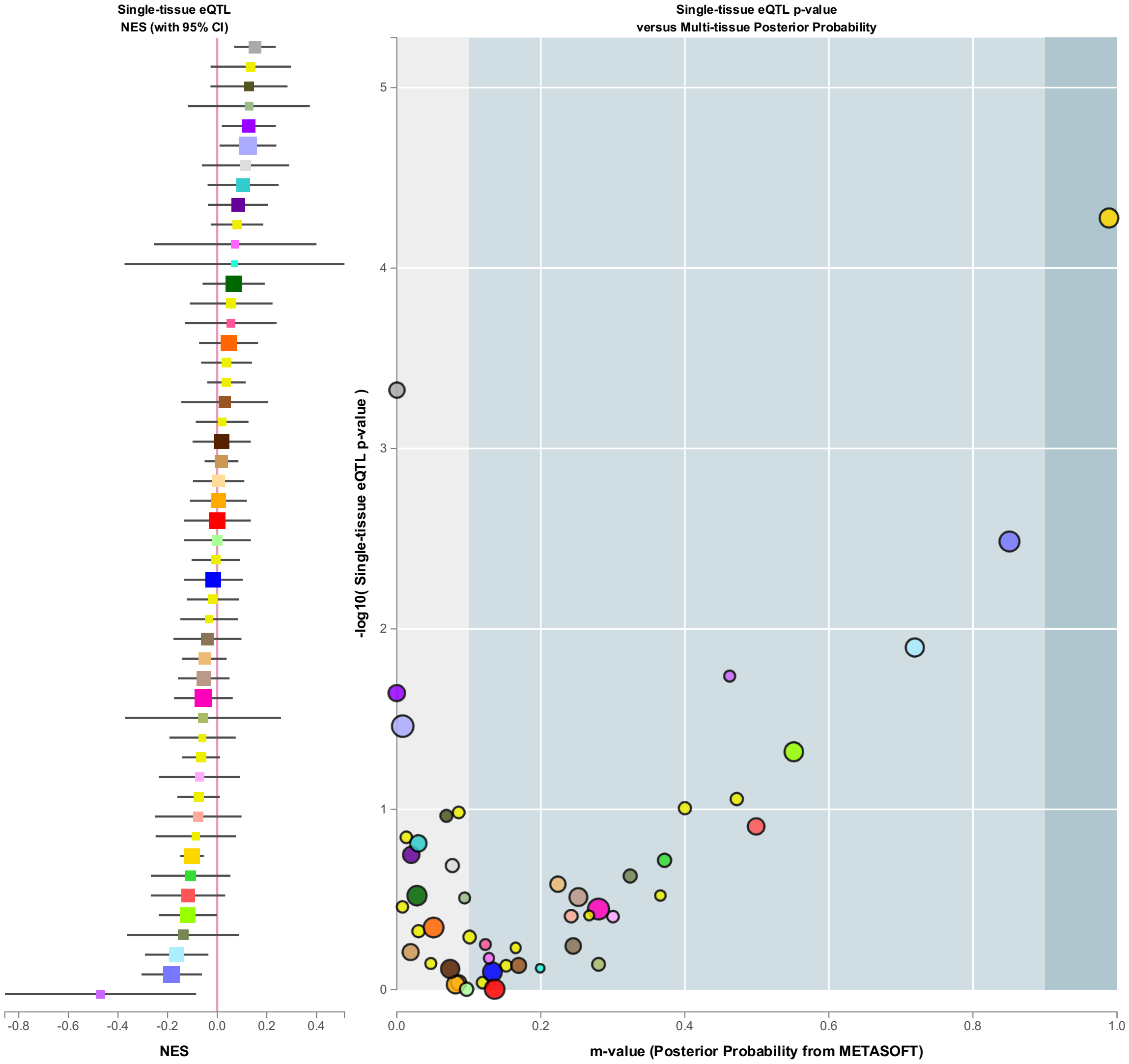


Supplemental Figure 5: **Multi-tissue eQTL Comparison. ENSG00000135472.8 FAIM2and chr12:49895449:G:A.** Image generated in https://www.gtexportal.org/home/snp/rs632151#eqtl-block
